# Multi-ancestry meta-analysis and fine-mapping in Alzheimer’s Disease

**DOI:** 10.1101/2022.08.04.22278442

**Authors:** Julie Lake, Caroline Warly Solsberg, Jonggeol Jeffrey Kim, Juliana Acosta-Uribe, Mary B. Makarious, Zizheng Li, Kristin Levine, Peter Heutink, Chelsea Alvarado, Dan Vitale, Sarang Kang, Jungsoo Gim, Kun Ho Lee, Stefanie D. Pina-Escudero, Luigi Ferrucci, Andrew B. Singleton, Cornelis Blauwendraat, Mike A. Nalls, Jennifer S. Yokoyama, Hampton L. Leonard

## Abstract

Genome-wide association studies (GWAS) of Alzheimer’s disease are predominantly carried out in European ancestry individuals despite the known variation in genetic architecture and disease prevalence across global populations. We leveraged published and *de novo* GWAS from European, East Asian, African American, and Caribbean Hispanic populations to perform the largest multi-ancestry GWAS meta-analysis of Alzheimer’s disease to date. This method allowed us to identify two independent novel disease-associated loci on chromosome 3. We also leveraged diverse haplotype structures to fine-map nine loci and globally assessed the heterogeneity of known risk factors across populations. Additionally, we compared the generalizability of multi-ancestry- and single-ancestry-derived polygenic risk scores in a three-way admixed Colombian population. Our findings highlight the importance of multi-ancestry representation in uncovering and understanding putative factors that contribute to Alzheimer’s disease risk.

## INTRODUCTION

Alzheimer’s disease (AD) is a complex genetic disorder with a range of deleterious variants across multiple genes attributed to both early and late-onset forms of sporadic AD ^1^. The strongest genetic risk factor for late-onset AD is *APOE*-*e4*, yet it has been estimated that there may be anywhere from 100 to 11,000 variants that also contribute to risk of late-onset AD ^2,3^. Large-scale genome-wide association studies (GWAS) in European ancestry populations have identified over 75 loci that are associated with AD and related dementias (ADD) ^4^. However, genetic research in AD that focuses solely on European populations limits additional discoveries afforded by studying diverse cohorts. Including non-European populations in genetic research provides new opportunities to uncover ancestry-specific risk variants and loci, increase statistical discovery power, improve fine-mapping resolution to identify putative causal variants, and identify loci with heterogeneous effects across ancestry groups ^5–7^.

Implementing existing ancestry-aware or heterogeneity penalizing meta-regression approaches have proven powerful at deconvoluting the genetic architecture of other phenotypes across populations ^8–18^. Here we report the results of a multi-ancestry genome-wide meta-analysis of the largest publicly available AD GWAS from individuals of European, East Asian, and African American ancestry, and a *de novo* GWAS of Caribbean Hispanic individuals. Using a meta-regression approach implemented in MR-MEGA, we demonstrate improved fine-mapping at several known AD loci and estimate the extent to which heterogeneity at these loci is attributable to genetic ancestry. This study highlights the utility of multi-ancestry analyses to further our understanding of disease biology and reduce health disparities in research by nominating novel loci and characterizing genetic differences across populations.

## RESULTS

### Data included in this study

Our multi-ancestry meta-analysis included a total of 54,233 AD cases, 46,828 proxy AD and related dementia (proxy-ADD) cases, and 543,127 controls (**Figure 1; Table S1**). Detailed information about the existing GWAS summary statistics used in this report are described elsewhere ^4,6,19^. In brief, the most recent publicly available AD GWAS includes 39,106 clinically diagnosed AD cases, 46,828 proxy-ADD cases (defined as having a parent with AD/dementia) and 401,577 controls of European ancestry ^4^. FinnGen data from Release 6 includes 7,329 AD cases and 131,102 controls free of any neurological disorder. We also included the largest publicly available AD GWAS of African American (2,748 cases and 5,222 controls) ^6^ and East Asian (3,962 cases and 4,074 controls) ^19^ populations and an additional *de novo* GWAS including 1,095 cases and 1,179 controls of Caribbean Hispanic ancestry. Select SNPs from the Gwangju Alzheimer’s & Related Dementias (GARD) East Asian cohort (1,119 cases and 1,172 controls) were used to assess East Asian risk at our novel loci post-hoc since these SNPs were not tested in the East Asian dataset used in our meta-GWAS ^20^. In this study, we considered significant variants as passing the standard p-value threshold of 5 × 10^−8^, consistent with most GWAS meta-analyses and used previously in other multi-ancestry studies ^21–23^. Our analysis included only variants that passed quality control and with a minor allele frequency > 1% in a minimum of three datasets to accurately quantify heterogeneity, effectively reducing the number of potential haplotypes and tests.

**Fig. 1:**
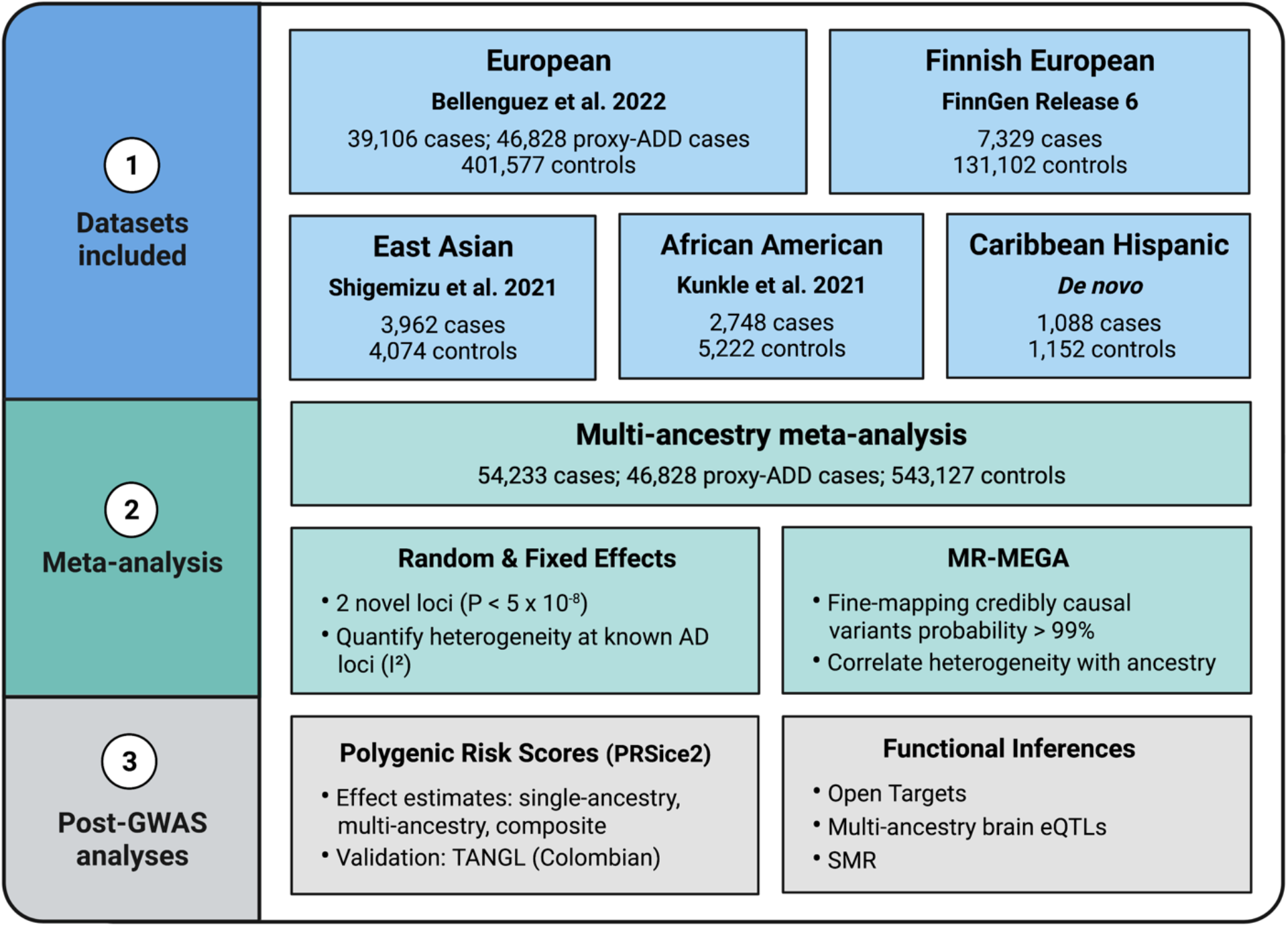
Outline of multi-ancestry meta-analysis procedure and downstream analysis. Created with BioRender.com.

### Meta-GWAS

Association summary statistics from all five datasets, representing four super populations, were aggregated via fixed and random effects models implemented in PLINK v1.9 ^24^ and a multi-ancestry meta-regression implemented in MR-MEGA ^25^ (**Table S1**). We did not observe any genomic inflation after excluding rare variants (MAF < 1% per study) and correcting for case-control imbalance (**Table S1; Figure S1**). Chromosome 19 was also excluded from genomic inflation estimates to avoid bias from the *APOE* region. Association results from the random effects and MR-MEGA meta-analyses were moderately concordant for SNPs without heterogeneity (I^2^=0, R^2^=0.6). Our study also demonstrated that MR-MEGA is advantageous for SNPs with heterogeneous allelic effects (**Figure S2**).

We replicated 51 known AD loci in the random effects and MR-MEGA meta-analyses, five of which only reached genome-wide significance (P < 5 × 10^−8^) using MR-MEGA and six of which were only significant using the random effects model (**Table S5**). We additionally identified two independent AD risk loci on chromosome 3 near *TRANK1* (rs9867455; P_RE_=3.49E-08, β_RE_=-0.0424, I^2^=0) and *VWA5B2* (rs9837978; P_RE_=3.75E-08, β_RE_=-0.0526, I^2^=0) that are outside of the maximal linkage disequilibrium (LD) boundary for any known AD risk loci (**Figure 2**). Both loci were identified using the random effects model and we did not identify any additional novel loci using MR-MEGA (**Figure 3a**). *TRANK1* is also a risk gene for bipolar disorder I (BD I) ^26^ and schizophrenia ^27^, although LocusCompare plots show a weak correlation with BD I (R^2^=0.46) and schizophrenia (R^2^=0.23) GWAS at this locus (**Figure S3-4**). We observed moderate LD (R^2^=0.44, 1000 Genomes EUR) between the lead SNP identified in our study (rs9867455) and the lead SNP identified in the BD I GWAS (rs9834970), indicating potential cross-disease overlap for this locus.

**Fig. 2:**
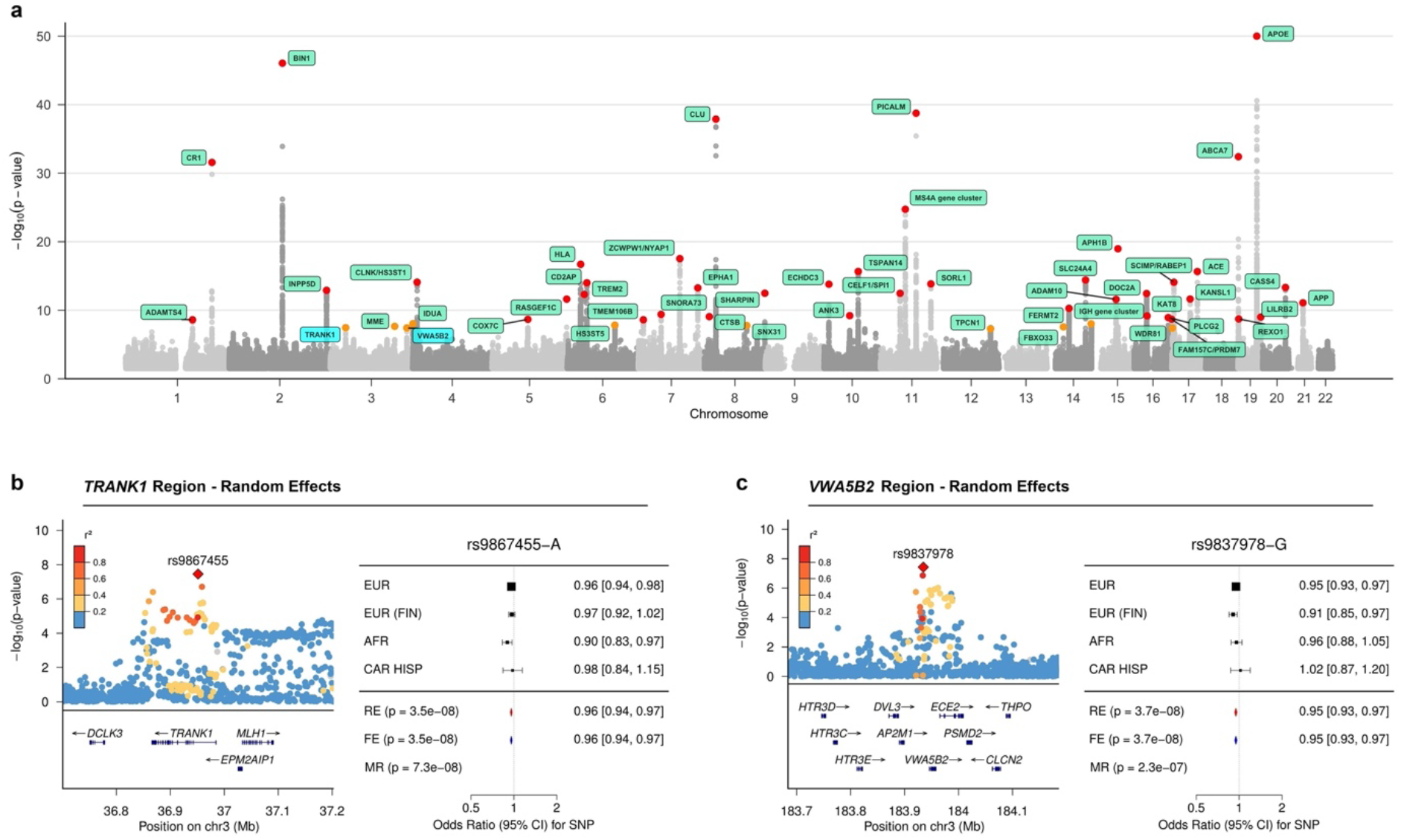
Summary of multi-ancestry meta-analysis. Panel A shows the Manhattan plot for the random effects meta-analysis P-values, truncated at -log_10_(P) < 50. Panels B and C show the corresponding local association plots for the two loci of interest. Panels C and D show forest plots summarizing the effect estimates per ancestry group for lead SNPs at the two loci of interest. Lead SNPs from both novel loci were absent in the East Asian dataset used for discovery. Abbreviations - MR: MR-MEGA, RE: Random effects, FE: Fixed effects, EUR: European, EUR (FIN): Finnish European, AFR: African American, CAR HISP: Caribbean Hispanic.

**Fig. 3:**
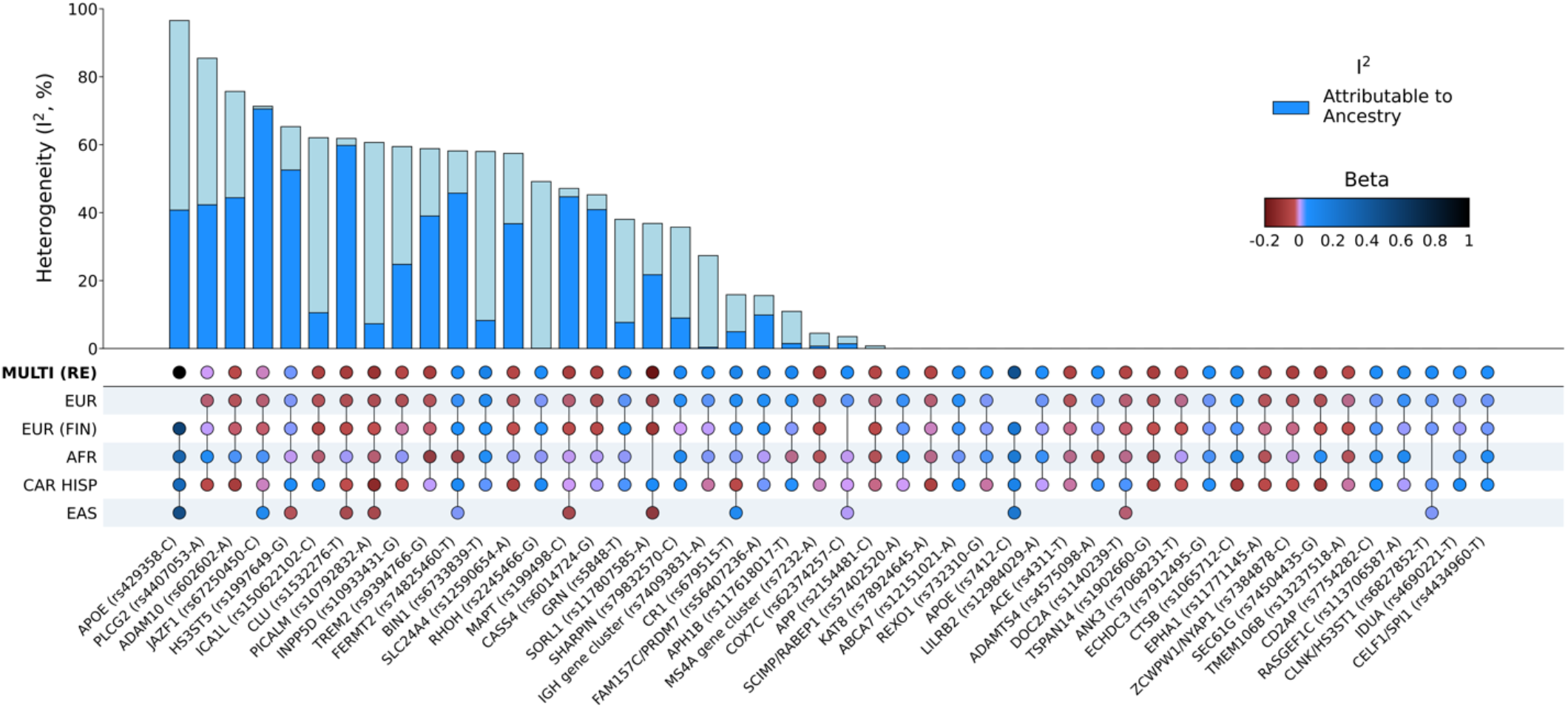
Graphical summary of heterogeneity at AD genetic risk loci. Lead SNPs were derived from MR-MEGA using maximal LD blocks, apart from *APOE* rs429358 and rs7412. Both *APOE* SNPs were absent in summary statistics from the most recent European-ancestry AD GWAS. Aggregate effects were estimated using a random effects model since MR-MEGA assumes that effects differ across populations. Allelic effect heterogeneity that is attributable to genetic ancestry was estimated using Cochran’s Q statistics for ancestral and residual heterogeneity from the meta-regression (**Online Methods**).

Since the lead SNPs for these potential novel loci were absent in the East Asian dataset used for initial discovery ^19^, we attempted to test these SNPs in an independent East Asian ancestry dataset ^20^. We observed a nominally significant association at *VWA5B2*-rs9837978 (P=0.048, β=0.204) in the GARD cohort (n=2,291), although the direction of effect was not consistent with the datasets included in the discovery GWAS. We were unable to test the association at *TRANK1*-rs9867455 since this SNP was not included in that GWAS and an LD proxy SNP was not available.

### Gene prioritization for novel loci

Using public expression quantitative trait locus (eQTL) evidence from Open Targets ^28^ and multi-ancestry brain eQTL summary data ^29^, we assessed whether *TRANK1-*rs9867455 and *VWA5B2*-rs9837978 are associated with the expression of nearby genes. Open Targets reported rs9867455 as a significant eQTL (P < 1 × 10^−6^) for *LRRFIP2, ITGA9, GOLGA4, MLH1*, and *TRANK1* across blood or other tissues. *LRRFIP2, GOLGA4* and *TRANK1* were also nominated in the multi-ancestry brain eQTL data. Open Targets reported rs9837978 as a significant eQTL for *AP2M1, ABCF3, VWA5B2, ALG3, ABCC5, DVL3*, and *CLCN2. AP2M1*, as well as two additional genes (*EIF2B5* and *ECE2*) were nominated in the multi-ancestry brain eQTL data.

To prioritize susceptibility genes with expression effects on AD risk, we performed summary-based Mendelian Randomization (SMR) to infer whether expression of the eQTL-nominated genes is causal for AD. More details regarding the purpose and methods used to perform SMR can be found in the Functional Inferences section of the **Online Methods**. At the *TRANK1-* rs9867455 locus, *TRANK1, LRRFIP2, GOLGA4*, and *ITGA9* were significant in our SMR results for affecting AD risk via expression across multiple tissue types. The strongest associations in cortex tissue were seen with *TRANK1* and *LRRFIP2*. The GWAS signal at the *TRANK1-* rs9867455 locus colocalized most strongly with *TRANK1* expression in cortex tissue (R^2^=0.52; **Figure S5**). At the *VWA5B2-*rs9837978 locus, *VWA5B2, AP2M1, ABCF3, ALG3, EIF2B5, DVL3, CLCN2, ABCC5* were significant in our SMR results. The strongest associations in brain tissues were seen in *ABCF3, ALG3* and *EIF2B5*, although colocalization between the GWAS signal and these eQTLS were not very strong (R^2^ < 0.5; **Figure S6**). For more details on directionality of these associations, see **Table S2**.

### Fine-mapping

A total of nine loci outside of the *APOE, MAPT*, and major histocompatibility complex (MHC) regions were fine-mapped to a credible set of ≤ 2 SNPs with a combined posterior probability (PP) of 99% (**Table 2; Figures S7-8**). The *MHC* and *MAPT* regions were excluded from fine-mapping due to a complex haplotype structure across populations ^30,31^ and known haplotype inversions ^32^, respectively. Five of these loci were previously fine-mapped with PP > 0.8 in large GWAS of European populations (**Figure S7;** *BIN1*-rs6733839; *INPP5D*-rs10933431; *ECHDC3*-rs7912495; *APH1B*-rs117618017; *ABCA7*-rs12151021) ^33,34^. Four additional AD loci with 1-2 variants in their 99% credible sets have not been previously fine-mapped (**Figure S8a-b**,**d-e**; *RHOH*-rs2245466; *CTSB*-rs1065712; *FAM157C/PRDM7*-rs56407236; *GRN*-rs5848). Interestingly, *GRN* and *CTSB* are also known risk loci for Parkinson’s disease ^35^, but LocusCompare plots show low genetic correlation at these loci (*GRN*, R^2^=0.36; *CTSB*, R^2^=0.0048) which may indicate that distinct causal variants drive the associations (**Figure S9**). One additional locus with a credible set size > 2 was fine-mapped to a single SNP with PP > 0.8 (**Figure S8c**; *SLC24A4*-rs12590654, PP=0.91, n=32 in 99% credible set). In addition, two SNPs with a PP ≥ 0.3 were annotated as missense variants (**Figure S10;** *MS4A6A*-rs7232, PP=0.54, n=4 in 99% credible set; *SHARPIN*-rs34674752, PP=0.30, n=5 in 99% credible set). Notably, our fine-mapping analysis did not replicate *SORL1*-rs11218343, which has been previously fine-mapped with a PP > 0.999 in two large European studies ^33,34^, likely due to a different regional architecture in the East Asian population as has been previously reported (**Figure S11**) ^36^. All 99% credible sets are provided in **Table S3**.

**Table 1:** Summary of novel loci.

**Table 2:** Fine-mapping results for all SNPs with a posterior probability (PP) > 0.8. Fine-mapped SNPs were considered novel if they were not previously fine-mapped with PP > 0.8 in two recent European-focused studies, Schwartzentruber et al. 2021 and Wightman et al. 2021.

### Heterogeneity analysis

We observed significant heterogeneity (I^2^ > 30%) at 19 of the 48 loci that reached genome-wide significance (P < 5 × 10^−8^) in MR-MEGA (**Figure 3**). At least 50% of the observed heterogeneity was attributable to genetic ancestry at 10 of these loci, of which four showed the strongest effect in East Asians (*SORL1, MAPT, JAZF1*, and *CLU*), four in Caribbean Hispanics (*FERMT2, SLC24A4, ADAM10*, and *HS3ST5*), one in non-Finnish Europeans (*CASS4*), and one in African Americans (*TREM2*) (**Figure 3, Figure S12**). Interestingly, four of these loci (*SLC24A4, FERMT2, CLU*, and *ADAM10*) showed an opposite direction of effect among African Americans compared to all other ancestry groups tested. We also assessed heterogeneity at lead SNPs from the most recent European GWAS ^4^ and found that 37% of the lead SNPs tested presented significant heterogeneity (I^2^ > 30%), of which 48% were primarily attributable to ancestry (**Table S4**). Five of the fine-mapped SNPs also showed significant heterogeneity (I^2^ > 30%), of which only *SLC24A4* showed heterogeneity that was primarily attributable to genetic ancestry (**Table 2**).

The genetic polymorphisms rs7412 and rs429358 that form the *APOE* e2/e3/e4 alleles presented very different allelic heterogeneity. Consistent with previous studies ^7,37^, we observed an attenuated signal at *APOE*-rs429358, which determines the *APOE*-e4 allele, among the cohorts of African Americans and Caribbean Hispanics (**Figure S13a**). *APOE*-rs429358 had the highest heterogeneity (I^2^=96.54) of the SNPs tested with ∼42% attributable to genetic ancestry, although these polymorphisms were not available to test in the most recent European-ancestry AD GWAS^4^. In contrast, *APOE*-rs7412, which determines the *APOE*-e2 allele, did not present any heterogeneity (**Figure S13b;** I^2^=0). Complementary to standard GWAS association tests, we also generated P-values representing heterogeneity of effect estimates attributable to genetic ancestry in the multi-ancestry meta-regression and observed the strongest signal near *APOE* (**Figure S14**). Additionally, we observed strong evidence of ancestry-related heterogeneity (P_HET_ < 1e-6) near *SORL1*, as well as *PAPOLG, AC026202*.*5*, and *snoU13* which did not meet genome-wide significance in the association results. LocusZoom and beta-beta plots of these loci suggest that non-European populations primarily drive these association signals, and there are likely discordant effects across populations (**Figure S15**).

### Polygenic risk scoring

We tested the performance of the multi-ancestry random effects model and each of the GWAS from single ancestral populations in a Colombian cohort of AD cases (n=281) and neurologically normal controls (n=87). This cohort is an admixture of three ancestral populations, with European substructure making up the highest proportion of global ancestry (mean of 64%, SD=15%), followed by Indigenous American (mean of 27%, SD=11%), and African (mean of 9%, SD=11%). Latino ancestry samples were used to test PRS applicability as they were a population not represented in the meta-analyses. Single-ancestry PRS performed worse than multi-ancestry random-effects-derived PRS in terms of area under a receiver operating characteristic curve (AUC), with maximal AUCs of 79% and 68% including and then excluding *APOE* variants in this population. Non-European AUCs tended to improve with increasing sample size (**Figure 4**), suggesting that the composite score, combining ancestry-specific PRS by population weights, may have performed as well or better than the random-effects-derived PRS if the component GWASs from under-represented populations were better powered.

**Fig. 4:**
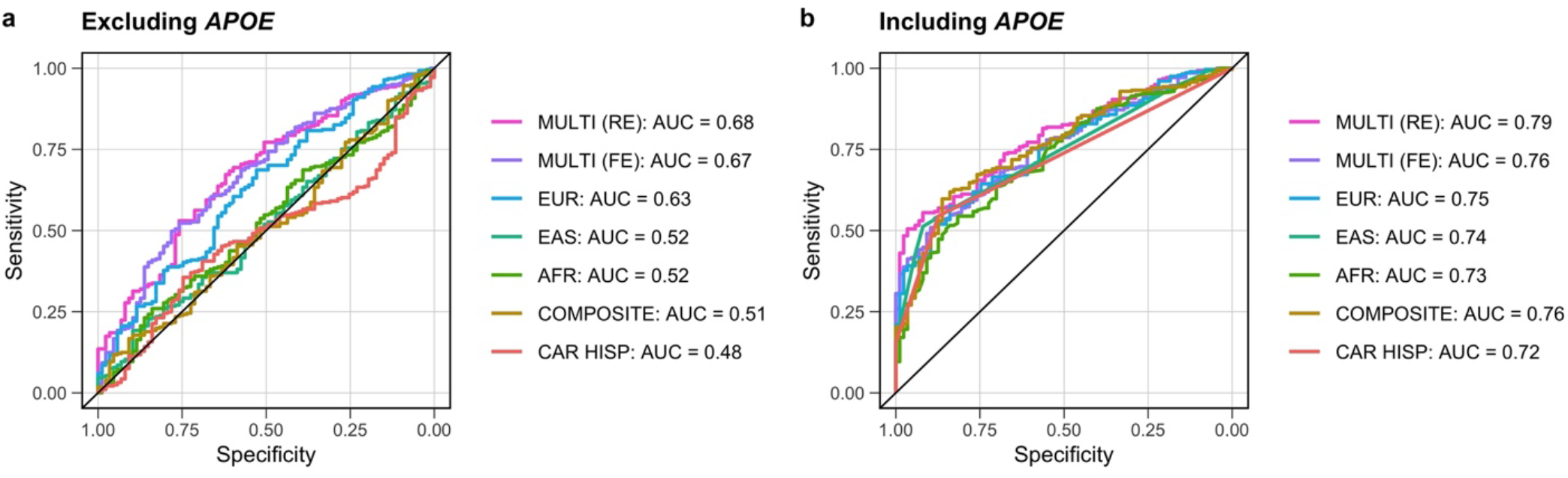
Graphical summary of genetic risk scores. These genetic risk scores were derived from multi-ancestry and ancestry-specific risk estimates, then applied to an admixed Colombian cohort to evaluate significance and predictive power. Panels A and B show the AUC for each genetic risk score with color coding to delineate the source of the risk estimates for scores excluding and then including *APOE-e4* variants.

## DISCUSSION

We performed a large, genome-wide meta-analysis of AD across five datasets, representing four super-populations. By leveraging data from multiple ancestry groups, we replicated 51 known AD loci and identified two novel risk loci on chromosome 3. The first is a signal near *TRANK1*, which encodes tetratricopeptide repeat and ankyrin repeat containing 1. *TRANK1* is associated with DNA- and ATP-binding and DNA repair and is highly expressed in brain tissue and female reproductive tissues ^38,39^. Previously, *TRANK1* has been cited as a robust risk locus for BD in European ^40,41^, East Asian ^42^, and multi-ancestry ^43^ studies, although subtype analyses suggest that this signal is primarily driven by the most heritable subtype, BD I. Previous SMR analysis of the *TRANK1* region in BD suggested that both *TRANK1* and *GOLGA4* may be susceptibility genes ^42^. *TRANK1* is also a risk locus for schizophrenia, which is genetically correlated with BD I^26^. BD has also been shown to increase risk for AD, with the two sharing significant genetic overlap ^44^. Previous studies have suggested that *TRANK1* may be involved in neural differentiation, blood brain barrier permeability changes, and neuroinflammation, all of which may be relevant to neurodegeneration ^42,45^. Interestingly, Kunkle et al. nominated *TRANK1* through a gene-based analysis conducted in an African American population, but not through the single SNP association testing included in this study ^6^. A combination of eQTL and SMR nominated *TRANK1, LRRFIP2, GOLGA4*, and *ITGA9* as potential genes underlying this SNP association, with the strongest associations in cortex tissue seen with *TRANK1* and *LRRFIP2* (**Table S2**). *LRRFIP2* encodes LRR binding FLII interacting protein 2, which regulates Toll-like receptor 4 (TLR4) and can downregulate the *NLRP3* inflammasome. TLR4 can induce microglial amyloid-β clearance in the brain in early stages of AD but can later induce an inflammatory response, suggesting that disruptions to *LRRFIP2* may affect AD pathology in patients ^46^.

The second locus is nearest to *VWA5B2*, which encodes von Willebrand factor A domain-containing protein 5B1. Von Willebrand factor (VWF) is a glycoprotein that facilitates blood clotting at areas of injury. High VWF is associated with short-term risk of dementia, possibly due to the increased risk of blood clots restricting blood flow in the brain ^47^. VWF can be elevated in response to inflammation, but *VWA5B2* was found to be downregulated in AD patients. Whether *VWA5B2* has biological implications on risk for AD needs to be further investigated ^48^. The lead variant rs9837978 does not lie within any of the nearby genes at this locus, but eQTL and SMR evidence for this variant nominated eight nearby genes including *VWA5B2* (**Table S2**). The strongest SMR association in brain tissue is seen with *ABCF3*. The ABCF3 protein is a member of the ATP-binding cassette (ABC) superfamily, which transport a variety of substrates across intra and extracellular barriers ^49^. Members of the ABC A subfamily, such as ABCA7 and ABCA1, have previously been nominated as AD risk genes ^4^. *ABCF3* is a unique family member in that it lacks a transmembrane domain but has been nominated as a candidate of TLR signaling, similar to *LRRFIP2* ^50^. In addition to inducing inflammatory responses, TLRs can affect microglial activity, synaptic plasticity, and tau phosphorylation, providing additional evidence to their importance in AD pathology ^51^.

Future studies will be required to further disentangle the potential roles of the nominated genes in the context of AD risk. The disparity seen at points between the results on Open Targets, which consists of largely European data, and the multi-ancestry eQTL results for nominated genes also highlights the need for more multi-modal reference data including diverse ancestries. However, it is also possible that there could be different mechanisms underlying disease risk conferred by the implicated loci across different populations.

Our study highlights the utility of multi-ancestry datasets at uncovering putative mechanisms that contribute to AD. Fine-mapping at several known AD loci was better resolved using the multi-ancestry meta-regression compared to previous efforts in European populations. For example, fine-mapping near *RHOH, CTSB* and *FAM157C/PRDM7* nominated variants that are located in untranslated regions that were not well-resolved in European studies. Variants in the 3’UTR region can impact translation or protein stability, and transcription binding can be impacted by variants in the 5’UTR region. Additionally, *GRN*-rs5848 is associated with circulating progranulin levels and decreased *GRN* expression has been implicated in several neurodegenerative diseases, including AD and frontotemporal dementia ^52–54^. In contrast to previous studies in European populations, the *SORL1* locus was not resolved to a single putative causal SNP. Lead SNPs in both the European (*SORL1*-rs11218343) and East Asian (*SORL1*-rs117807585) GWAS are more common among East Asians compared to all other populations in the Genome Aggregation Database v2.1.1 (rs11218343: AF_EAS_=0.30, AF_EUR_=0.039; rs117807585: AF_EAS_=0.22, AF_EUR_=0.020). It is possible that alternative fine-mapping approaches that allow for multiple causal variants per locus will provide greater insight into the *SORL1* locus.

At the *MS4A* gene cluster, multi-ancestry fine-mapping resolved the signal to a credible set of five variants, with a common missense variant (rs7232, PP=0.54) and an intergenic variant nearest *MS4A4A* (rs1582763, PP=0.45) that are in moderate LD (R^2^=0.55, 1000 Genomes all populations) having the highest probability of causality (**Figure S10**). *MS4A4A* and/or *MS4A6A* modulate soluble TREM2 (sTREM2) in cerebrospinal fluid (CSF), which is correlated with AD progression. Previous studies have shown that rs7232 is associated with *MS4A6A* gene expression and CSF sTREM2 ^55,56^, while rs1582763 is a cis-eQTL for *MS4A4A* and *MS4A6A* ^57^. Conditional analysis of CSF sTREM2 levels in this region have pointed to two independent signals represented by rs1582763 and rs6591561 (MS4A4A p.M159V) ^57^. Therefore, a fine-mapping approach that allows for multiple causal variants may be more appropriate for this region.

In addition to highlighting genetic risk factors that are shared across populations, our results also highlight AD loci with significant heterogeneity that may reflect variation in effect sizes, allele frequencies or interaction(s) with environmental risk factors that vary by ancestral group. For example, we observed the strongest evidence of heterogeneity at *APOE*-rs429358. Around 42% of the heterogeneity at this allele was attributable to genetic ancestry, while the remaining heterogeneity may reflect other sources of variation such as imputation accuracy since this allele is rarely assayed successfully on genotyping arrays. At *JAZF1*-rs67250450 and *CLU*-rs1532276, we observed the strongest evidence of ancestry-related heterogeneity, both of which are most common among individuals of East Asian ancestry and showed the strongest effects in this population (**Figure 3**). We also observed significant ancestry-related heterogeneity at *SORL1* and *TREM2*, which have been previously shown to harbor population-specific risk variants ^58,59^. Given that our analyses focused on common variation, the effects of rare heterogeneous variants (e.g. *ABCA7*-rs115550680, which has comparable effects to *APOE*-rs429358 among African Americans ^60^) may not have been fully captured.

While this study marks progress towards assessing genetic risk of AD across multiple populations, we acknowledge several limitations. MR-MEGA is a useful tool for fine-mapping and ancestral heterogeneity estimation, but the software requirements of population overlap (K > 3) often result in reduced variant sets after study level quality control. This can bias fine-mapping results as we reduce the potential resolution on local haplotypes, and usually necessitates the inclusion of at least one of the larger European-focused studies. In our case, previous European and Finnish studies served as the backbone of our meta-GWAS. We did not replicate previous fine-mapping at *NCK2, TREM2* and *RNF223* from European-focused studies since study level quality control included filtering for common (MAF > 1%), biallelic SNVs due to potentially poor imputation and general low power for rare variants across ancestral groups. We acknowledge that variants with a minor allele frequency < 1% in one or more populations, as well as indels and structural variants, may contribute to the observed associations. Future work should include imputation using diverse reference panels from long read sequence data specific to AD to improve genomic coverage and provide insights into structural variation that may be population specific.

In addition, the number of axes of genetic variation (T) in MR-MEGA is restricted to T ≤ K-2, where K is the number of input GWAS. The East Asian GWAS used in our meta-GWAS tested less than half as many SNPs as the others (**Table S1**), limiting the meta-regression to a single axis of genetic variation (PC0) at SNPs that overlap the remaining GWAS (K=4). Including a larger number of input GWAS from underrepresented populations will likely improve the heterogeneity estimates outlined in this study.

While our study is inclusive, due to data availability and the European-dominated nature of genetic research, European-ancestry individuals make up approximately 85% of cases and the discovery efforts here maintain a baseline level of Eurocentric bias. Additionally, while our novel method of creating a composite PRS model that leverages admixture percentages is a potentially promising approach for assessing AD risk across ancestrally heterogeneous and/or admixed cohorts, its performance relies on sufficient sample sizes and global genetic representation. As larger scale GWAS for multiple continental “super populations” continue to become available, we believe this method of tuning PRS to an individual’s genetic admixture could have utility in a precision medicine context. Reducing the Eurocentric bias in AD genetics research will require the harmonization and refining of diagnosis in non-European research sites that serve communities with unique cultural and logistic concerns for participation in research. It is our hope to improve representation in AD genetic studies in the future, increasing the balance between European and well-powered non-European cohorts.

## ONLINE METHODS

### Existing GWAS studies

Summary statistics from Bellenguez et al. 2022 were accessed through the National Human Genome Research Institute-European Bioinformatics Institute GWAS catalog under accession number GCST90027158 in May 2022. Summary statistics from FinnGen Release 6 were accessed at https://www.finngen.fi/en/access_results in April of 2022. Summary statistics from Kunkle et al. 2021 were accessed through NIAGADS (https://www.niagads.org/) under accession number NG00100 in April of 2022. Summary statistics from Shigemizu et al. 2021 were accessed through the National Bioscience Database Center (NBDC) at the Japan Science and Technology Agency (JST) at https://humandbs.biosciencedbc.jp/en/ through accession number hum0237.v1.gwas.v1 in April of 2022. All summary statistics were aligned to GRCh37 and cleaned to remove indels, multi-allelics and rare variants (MAF < 1%) prior to multi-ancestry analysis.

### De-novo GWAS

#### Caribbean Hispanic GWAS

Data from the Columbia University Study of Caribbean Hispanics and Late Onset Alzheimer’s disease were accessed via application to dbGaP accession number phs000496.v1.p1 in April of 2022. Samples were filtered to keep unrelated individuals without missing values for AD affection status, age, study category, education, and a missing call rate < 0.02. Principal component analysis (PCA) was performed on a combined dataset of study subjects and HapMap was used as a reference to identify potential outliers. Controls with a family history of dementia were removed. Variant QC included exclusion filters for monomorphic SNPs, variants with MAF < 1%, missingness rates > 2%, sex differences in allelic frequency ≥ 0.2 and heterozygosity > 0.3, duplicate SNPs, Hardy–Weinberg Equilibrium (HWE) P-value < 1 × 10^−4^, and > 1 discordant calls or Mendelian errors. All variants with a significant frequency mismatch (χ^2^ > 300) with the TOPMed reference panel were removed prior to imputation.

Using PLINK v1.9 ^24^, we evaluated the association between AD and imputed genotypes via logistic regression on allele dosages with imputation quality > 0.3, adjusting for sex, age (age at disease onset for cases, age last seen for controls), education, study category, and the first 10 principal components (PCs). Study category denotes subcategories within the Caribbean Hispanic dataset (individuals are from the United States, Puerto Rico and the Dominican Republic) and is included to account for potential batch effects.

### Meta-analysis and fine-mapping

#### Meta-analysis

Three models were used to conduct multi-ancestry meta-analyses. Fixed effect and random effects models were performed using PLINK v1.9, while a separate analysis was performed using MR-MEGA v0.2 ^25^. MR-MEGA is well-powered to detect associations at loci with allelic heterogeneity since axes of genetic variation are included as covariates in the model. The European and Finnish European GWAS were included separately to account for finer-scale differences in allele frequencies. To determine the optimal number of PCs needed to distinguish cis- and trans-ancestry AD summary statistics, we visually inspected pairwise PC plots generated using all five GWAS referenced in **Table S1**. We observed adequate separation between the Caribbean Hispanic, European, African American, and East Asian GWAS using the first two meta-regression PCs (**Figure S16**). To increase the variant set, we also ran MR-MEGA separately for each combination of four input GWAS. A single axis of genetic variation (T=1) was included in this analysis since this is the maximum allowable given the constraints of the model (T ≤ K-2), where K is the number of input GWAS. Summary statistics were aggregated to maximize the effective sample size for each variant.

#### Fine-mapping

Fine-mapping was performed using approximate Bayes’ factors in favor of association from the meta-regression model implemented in MR-MEGA. In brief, FUMA was used to find maximal LD blocks around loci that reached P < 5 × 10^−8^ in the MR-MEGA analysis. LD blocks of independent significant SNPs (R^2^ >0.3, 1000 Genomes all populations) were merged into a single genomic locus if the distance between LD blocks was less than 250kb. Posterior probabilities (PP) were calculated using single-SNP Bayes factors and credible sets were generated for each locus until the cumulative PP exceeded 99%. All SNPs in the 99% credible sets were annotated with VEP (http://grch37.ensembl.org/Homo_sapiens/Tools/VEP) using default criteria to select one block of annotation per variant (**Table S3**).

### Assessment of allelic effect heterogeneity

Allelic effect heterogeneity between studies was assessed for all lead SNPs reaching genome-wide significance (P < 5 × 10^−8^) in the meta-regression, implemented in MR-MEGA. Genomic loci were defined as discussed in the fine-mapping methods. The meta-regression model derives axes of genetic variation from pairwise allele frequency differences between the input GWAS. Heterogeneity is then partitioned into (1) ancestry-related heterogeneity that is correlated with the axes of genetic variation and (2) residual heterogeneity that is likely due to other factors such as study design (e.g. covariate adjustments, phenotype definition, imputation quality) and/or geographical region. Total heterogeneity at each index SNP was quantified using the I^2^ statistic in PLINK v1.9 to avoid bias due to sample size for SNPs not tested in the large European studies. The I^2^ statistic describes the proportion of variation in effect estimates that is due to heterogeneity. We considered SNPs with an I^2^ > 30% as having significant heterogeneity since this suggests at least moderate variation in allelic effects ^61^. The percentage of this heterogeneity that is attributable to genetic ancestry was then calculated using Cochran’s Q statistics for ancestral and residual heterogeneity from the meta-regression (equation 1; ANC: ancestry, RESID: residual).

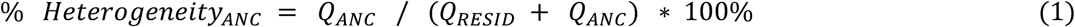

### Functional inferences

To prioritize genes underlying the two novel loci, we first looked at public eQTL data to determine whether the GWAS-identified lead variants are eQTLs for nearby genes. This allowed us to cast a wide net of potential regional genes of interest. We employed Open Targets for this effort, which shares eQTL results for variants from blood, brain, and a wide array of tissues from multiple public eQTL datasets ^28^. We additionally investigated a multi-ancestry brain eQTL dataset ^29^ which is currently not available on Open Targets. We considered the lead variants as significant eQTLs for a gene if they passed the significance threshold of P < 1 × 10^−6^, which has been shown to correspond to a genome-wide false discovery rate (FDR) of 5%, although we do acknowledge this may be overly conservative in our regional analyses ^62^.

Once we had nominated potential genes for which our lead variants were significant eQTLs, we used summary-based Mendelian Randomization (SMR) to make functional inferences as to whether the disease risk SNPs in these regions mediate gene expression. We integrated summary-level data from the most recent AD GWAS ^4^ with data from multiple eQTL studies in different tissues using the SMR method ^63^. SMR uses summary statistics to determine if an exposure is associated with a trait through a shared casual variant. MR can be used to mimic a randomized controlled trial, as having a variant that increases or decreases expression of a gene may be comparable to life-long treatment with a drug targeting the encoded protein of that gene.^64^ For example, if SNP A affects gene B expression (the exposure), and SNP A is also associated with AD risk (the outcome), you can infer the causal effect of the expression of gene B on AD risk.

We limited our results to the genes that were prioritized by our eQTL search and considered a gene significant for expression effect on a disease if it passed an FDR-adjusted SMR significance threshold of P < 0.05 and a HEIDI threshold of P > 0.01. Filtering for a HEIDI P-value of this magnitude helps to remove associations that are likely due to polygenicity and have violated the central assumptions of SMR. Finally, we assessed the colocalization between the SMR-nominated genes in brain tissues and the multi-ancestry random effects GWAS using LocusCompare ^65^.

### Polygenic risk scoring

#### PRS application cohort

Whole genomes from the Colombian population were accessed from “The Admixture and Neurodegeneration Genomic Landscape (TANGL) study and quality controlled as previously described ^58^. The TANGL cohort was further quality controlled in PLINK v1.9 to remove carriers of pathogenic variants for mendelian forms of dementia, as well as related individuals for a final cohort of 281 cases and 87 controls.

#### Pre-PRS variant alignment

Base summary statistics were pruned with the MungeSumStats R package ^66^ to remove multiallelic variants, align reference alleles to build GRCh37, and adjust weights for the appropriate reference alleles. The target TANGL cohort was also filtered to keep only bi-allelic variants and aligned to the same reference using PLINK v2.0.

#### PRS method

Polygenic risk score (PRS) analyses can be used to estimate an individual’s genetic liability to a phenotype by calculating the sum of risk allele effect size weights for an individual. Weights for the PRS were obtained from β estimates generated from multi-ancestry random and fixed effects meta-analyses as well as from individual ancestry summary statistics. PRS analyses were conducted using PRSice v2.3.5 including variants with minor allele frequency > 5%, genotype missingness < 10%, sample missingness < 10%, and HWE p-value < 1 × 10^−6^. The *APOE* region (with ranges defined by FUMA as described previously) was excluded prior to variant clumping. For PRS analyses including *APOE*, the genetic polymorphisms rs7412 and rs429358 were added to the QC’d summary statistics prior to variant clumping. β estimates for the APOE polymorphisms were not available in the Bellenguez et al. summary statistics ^4^ and therefore European-ancestry estimates were taken from another recent AD GWAS by Schwartzentruber et al. ^34^.

Variants were clumped in each 500 kb window with the index SNP at the center, an r^2^ threshold of 0.3, and a clump P-value threshold of 1. Sex, age, and the first 5 PCs were used as covariates in the PRS analysis. PCs were generated from non-imputed genotype data using FlashPCA ^67^. Variants with a MAF < 1%, genotype missingness < 10%, sample missingness < 10%, and HWE P-values < 5 × 10^−6^ were excluded using PLINK v1.9. The remaining variants were pruned with a 1000-kb window, a 10-SNP shift per window and an r^2^ threshold of 0.02 prior to PC calculation. PRS analysis was performed at select P-value thresholds to determine the best fit model (P=5 × 10^−10^, 5 × 10^−9^, 5 × 10^−8^, 5 × 10^−7^, 5 × 10^−6^, 5 × 10^−5^, 5 × 10^−4^, 5 × 10^−3^, 5 × 10^−2^). To assess the performance of each model, receiver operator characteristic curves were created using the pROC library in R for the best fit model from each analysis. An additional “composite” ROC curve was generated through a linear combination of each super population, with each PRS weighted by its associated admixture population percentage, previously determined in the TANGL cohort for each individual (equation 2; AFR: African American, EUR: European (including Finnish), EAS: East Asian, NAT: Native American) ^58^. Given the population history and similarities in haplotype structure between the East Asian and Native American populations, Native American admixture proportions were used to weight the East Asian PRS ^68,69^.

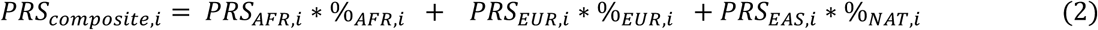

## Supporting information

Main Tables

## Data Availability

Code will be made available on the CARD GitHub after peer review: https://github.com/NIH-CARD/
Summary statistics from this study will be available to browse and download via our collaboration with the Broad's Neurodegenerative Disease Knowledge Portal after peer review: https://ndkp.hugeamp.org:8000/

## DATA AND CODE AVAILABILITY

Code is available on the CARD GitHub linked here.

Summary statistics from this study will be available to browse and download via our collaboration with the Broad’s Neurodegenerative Disease Knowledge Portal linked here after peer review.

## ACKNOWLEDGEMENTS

We want to acknowledge the participants and investigators of the FinnGen study. Assistance with phenotype harmonization and genotype cleaning, as well as with general study coordination, was provided by the Genetic Consortium for Late Onset Alzheimer’s Disease. Additionally, we want to thank Dr. Francisco Lopera and the Neuroscience Group of Antioquia and the University of Antioquia (Colombia) for letting us use the TANGL cohort.

## Funding acknowledgement

This work was supported in part by the Intramural Research Programs of the National Institute on Aging (NIA) part of the National Institutes of Health, Department of Health and Human Services; project numbers 1ZIA-NS003154, Z01-AG000949-02, ZO1-AG000535 and Z01-ES101986. J.S.Y. is supported by NIH-NIA R01 AG062588, R01 AG057234, P30 AG062422; NIH-NINDS U54 NS123985; the Rainwater Charitable Foundation; the Alzheimer’s Association; the Global Brain Health Institute; and the Mary Oakley Foundation. This research was supported by the Korea Brain Research Institute basic research program funded by the Ministry of Science and ICT (22-BR-03-05). Funding support for the “Genetic Consortium for Late Onset Alzheimer’s Disease” was provided through the Division of Neuroscience, NIA. The Genetic Consortium for Late Onset Alzheimer’s Disease includes a genome-wide association study funded as part of the Division of Neuroscience, NIA.

## AUTHOR INFORMATION

### Equal Contribution

**These authors contributed equally:** Julie Lake and Caroline Warly Solsberg as well as Jennifer Yokoyama and Hampton Leonard.

### Contributions

**Designed the study**.

Julie Lake, Caroline Warly Solsberg, Peter Heutink, Luigi Ferrucci, Andrew B. Singleton, Mike A. Nalls, Jennifer S. Yokoyama, and Hampton L. Leonard

**Acquired and processed data**.

Julie Lake, Caroline Warly Solsberg, Juliana Acosta-Uribe, Mary B. Makarious, Zizheng Li, Chelsea Alvarado, Dan Vitale, Sarang Kang, Jungsoo Gim, Stefanie D. Pina-Escudero, Luigi Ferrucci, Andrew B. Singleton, Cornelis Blauwendraat, Mike A. Nalls, Jennifer S. Yokoyama, and Hampton L. Leonard

**Analyzed data**.

Julie Lake, Caroline Warly Solsberg, Jonggeol Jeffrey Kim, Juliana Acosta-Uribe, Mary B. Makarious, Zizheng Li, Chelsea Alvarado, Dan Vitale, Sarang Kang, Jungsoo Gim, Stefanie D.

Pina-Escudero, Andrew B. Singleton, Cornelis Blauwendraat, Mike A. Nalls, Jennifer S. Yokoyama, and Hampton L. Leonard

**Interpreted data, writing and revising the first draft of the manuscript**.

All authors.

