## Supplementary Information for "Multi-ancestry meta-analysis and fine-mapping in Alzheimer’s Disease"

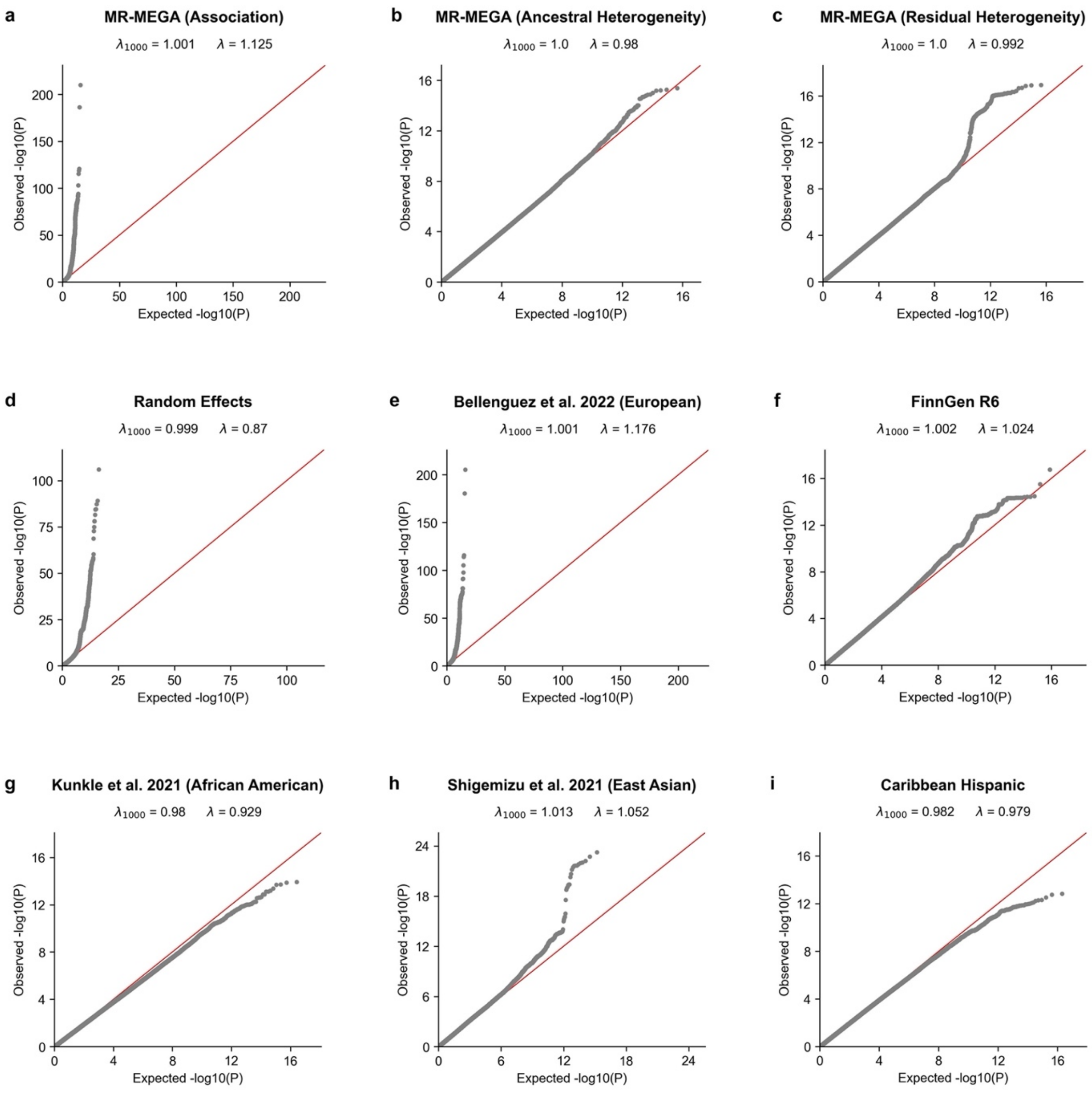


**Fig. S1**: Quantile-quantile plots and corresponding genomic inflation estimates, with and without scaling to 1000 cases and 1000 controls. MR-MEGA P-values are shown for association, ancestral heterogeneity, and residual heterogeneity. Chromosome 19 was excluded from all datasets to avoid bias by the *APOE* region. All summary statistics were filtered for MAF > 1%.

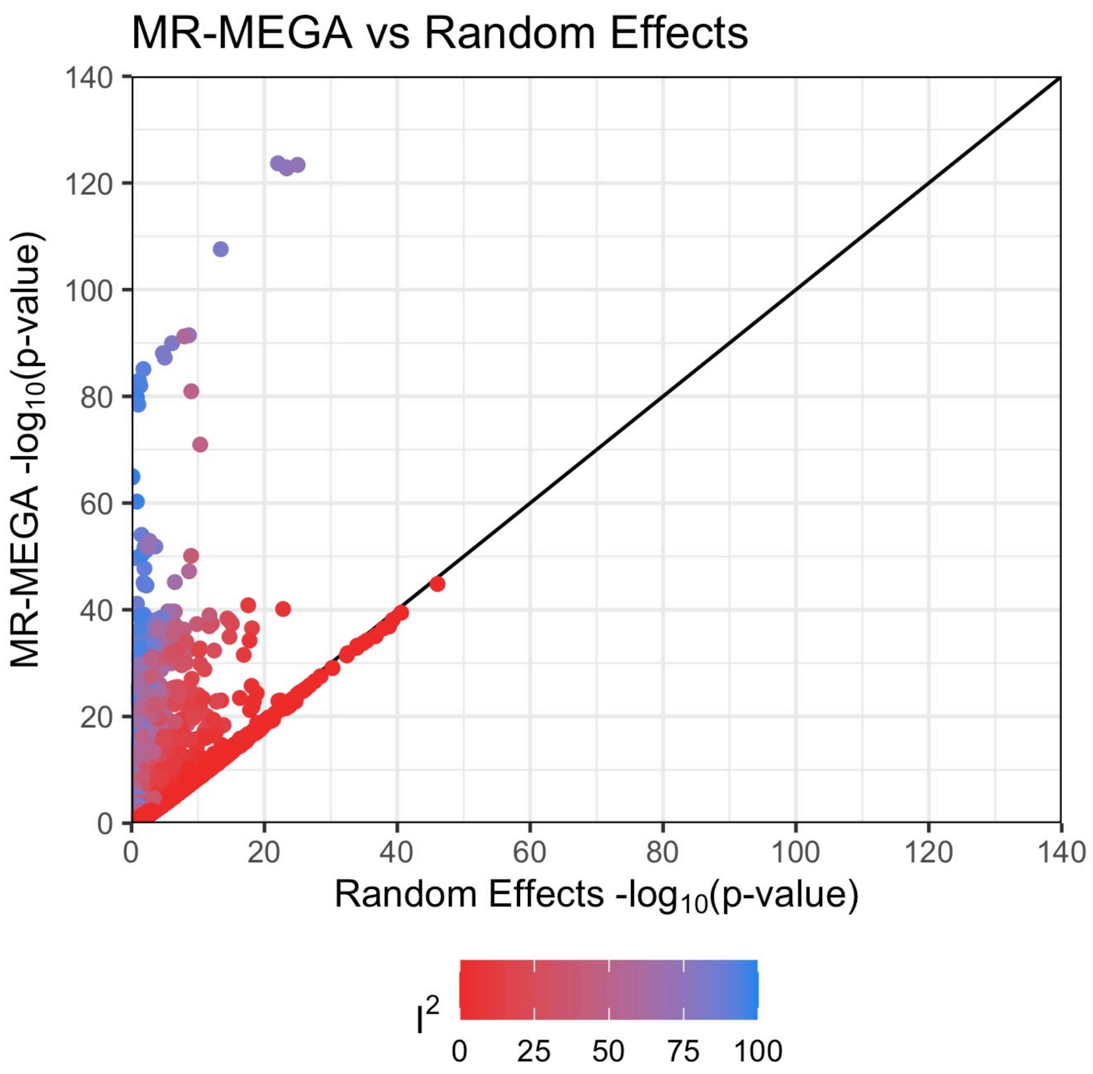


**Fig. S2:** MR-MEGA -$\log_{10}$ P-values plotted against random effects -$\log_{10}$ P-values. SNPs are colored by $I^2$ value and are limited to those present in at least 4 datasets.

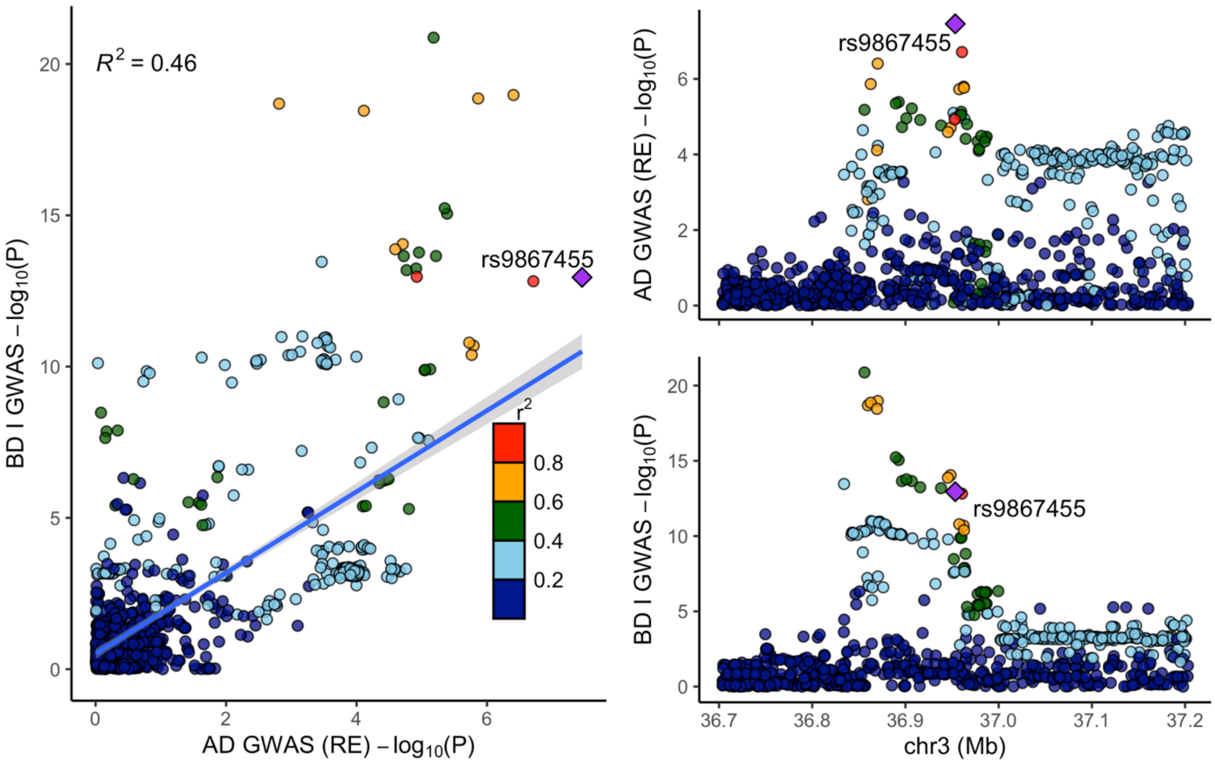


**Fig. S3:** LocusCompare plot for bipolar disorder I (BD I) and AD at the *TRANK1* locus. Reference LD patterns are based on the European population from 1000 Genomes. Points represent SNPs plotted at their -$\log_{10}$ P-values.

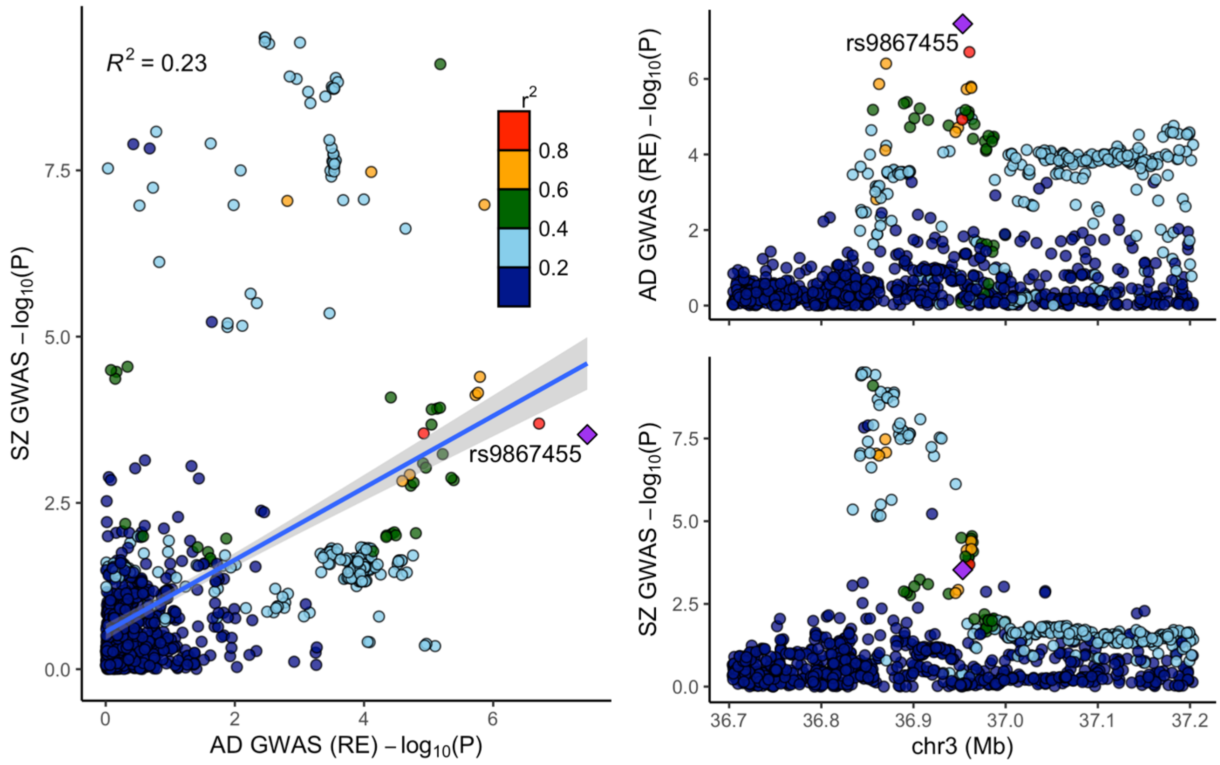


**Fig. S4:** LocusCompare plot for schizophrenia (SZ) and AD at the *TRANK1* locus. Reference LD patterns are based on the European population from 1000 Genomes. Points represent SNPs plotted at their -$\log_{10}$ P-values.

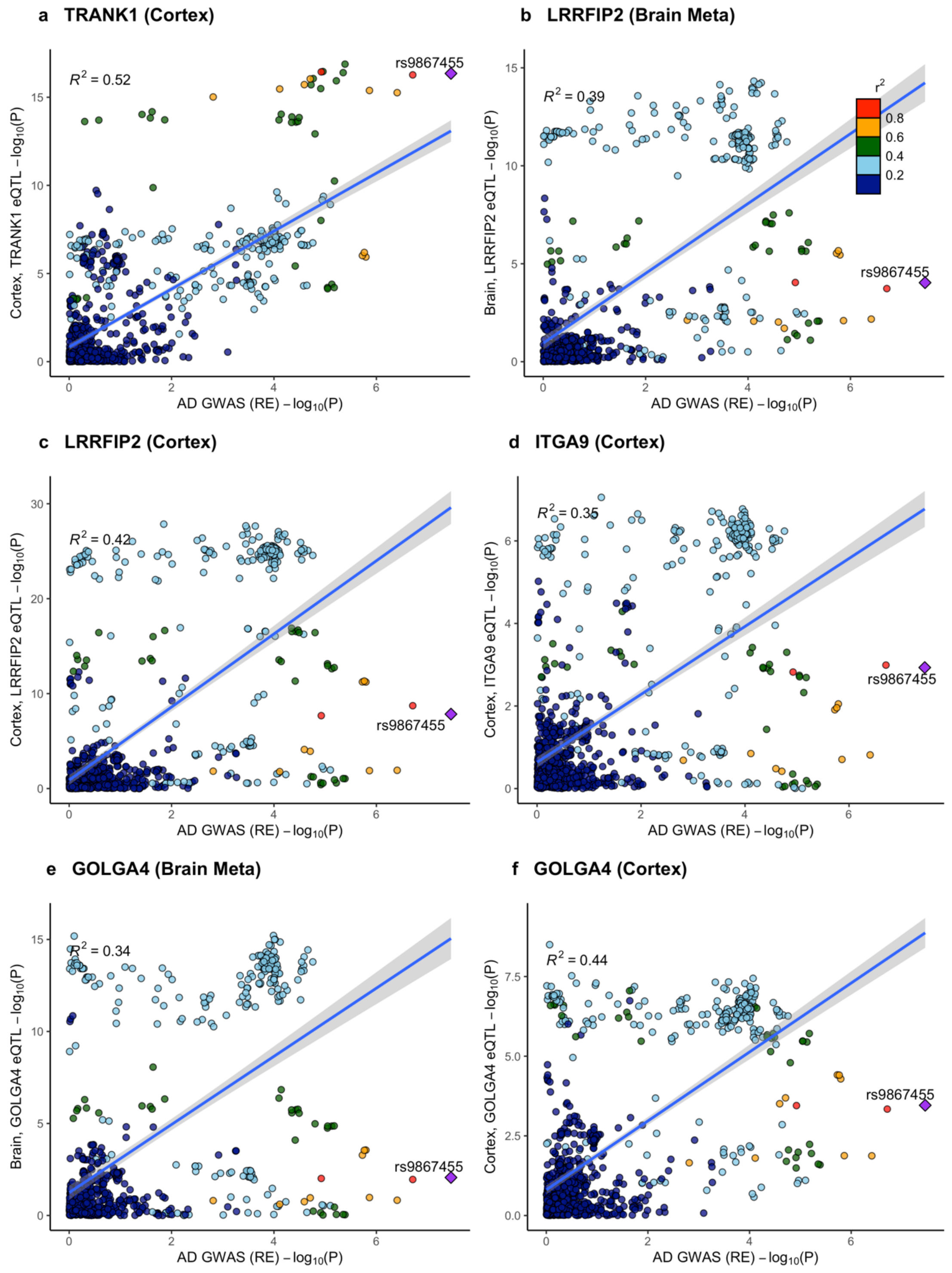


**Fig. S5:** LocusCompare plots showing colocalization at the *TRANK1* locus between the random effects meta-analysis results and brain eQTLs ($P < 1 \times 10^{-6}$) in genes that were significant in SMR (FDR $P < 0.05$). Reference LD patterns are based on the European population from 1000 Genomes. Points represent SNPs plotted at their $-\log_{10}$ P-values.

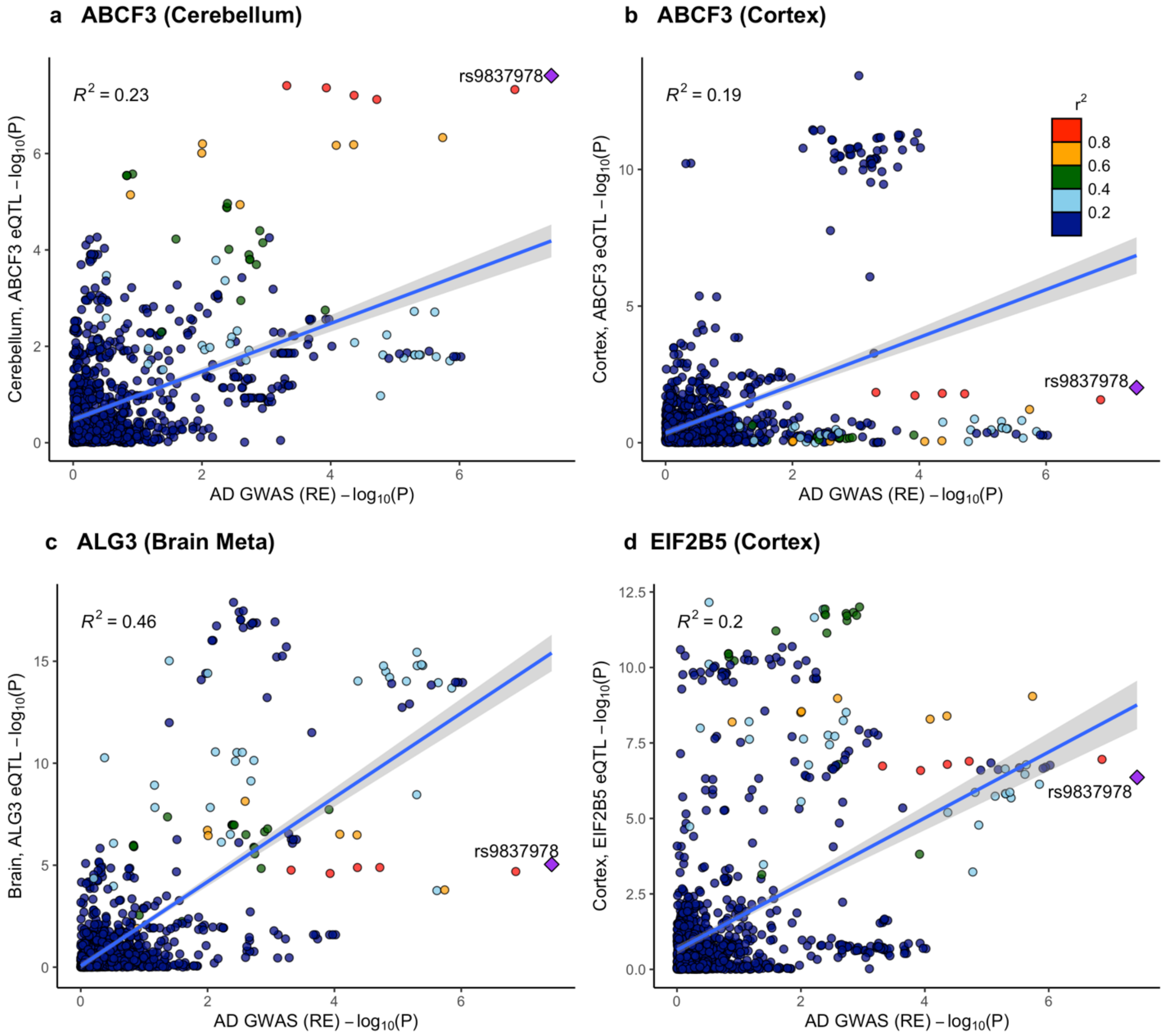


**Fig. S6:** LocusCompare plots showing colocalization at the *VWA5B2* locus between the random effects meta-analysis results and brain eQTLs ($P < 1 \times 10^{-6}$) in genes that were significant in SMR (FDR $P < 0.05$). Reference LD patterns are based on the European population from 1000 Genomes. Points represent SNPs plotted at their $-\log_{10}$ P-values.

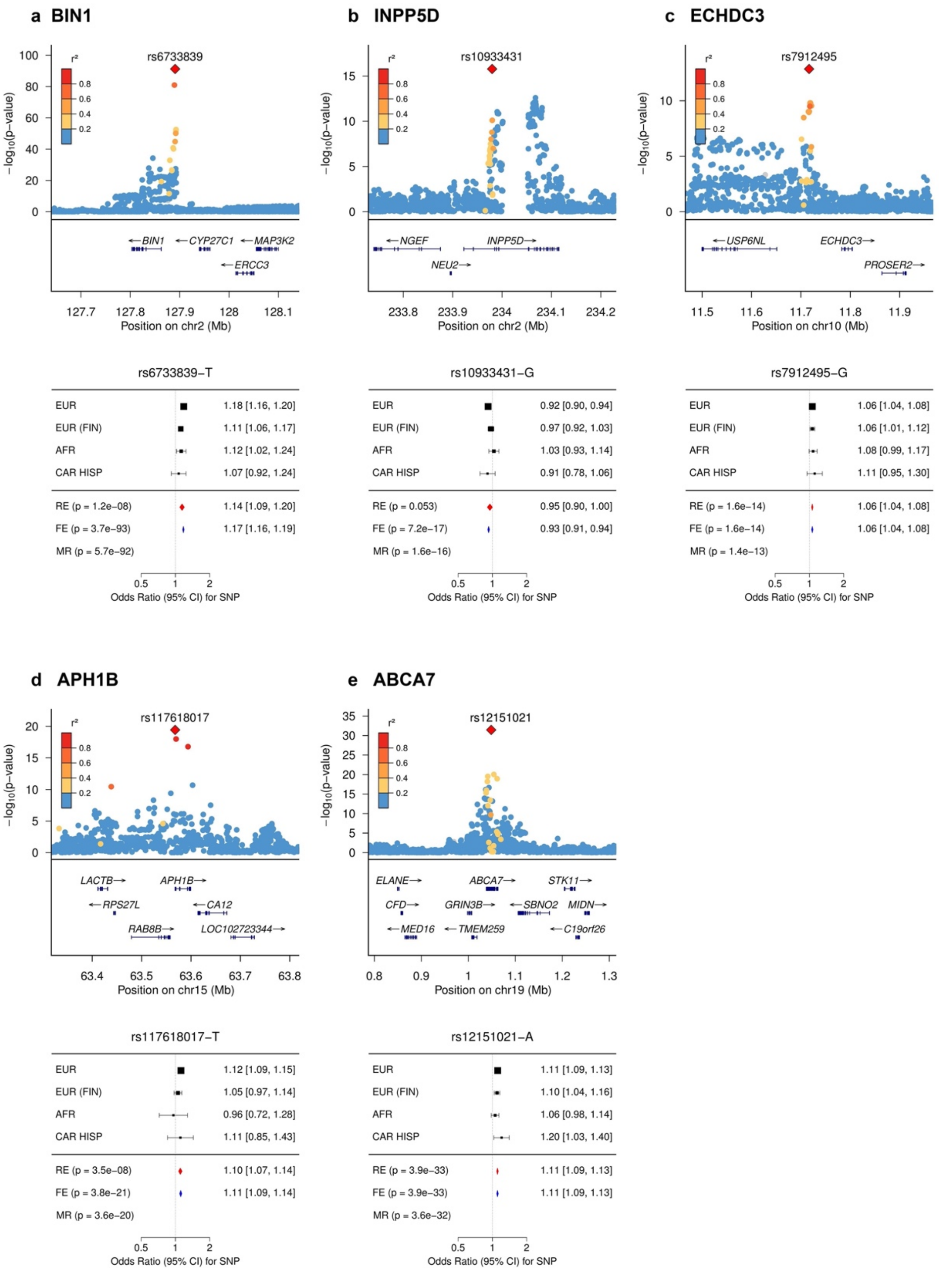


**Fig. S7:** LocusZoom and forest plots for SNPs fine-mapped in our study with posterior probability (PP) > 0.8 that have been previously fine-mapped in European studies. Reference LD patterns are based on all populations from 1000 Genomes.

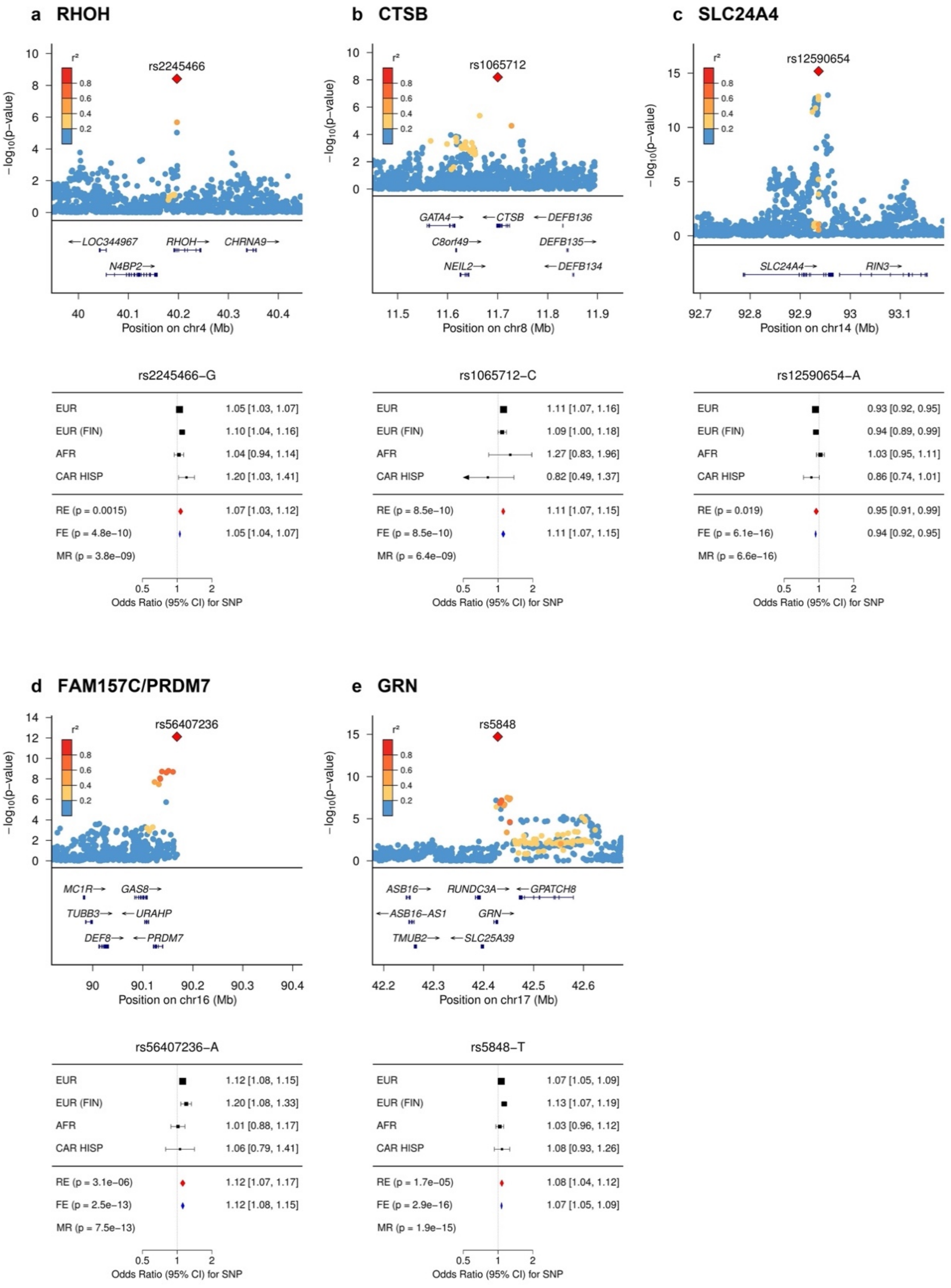


**Fig. S8:** LocusZoom and forest plots for SNPs fine-mapped in our study with posterior probability (PP) > 0.8 that have **not** been previously fine-mapped. Reference LD patterns are based on all populations from 1000 Genomes.

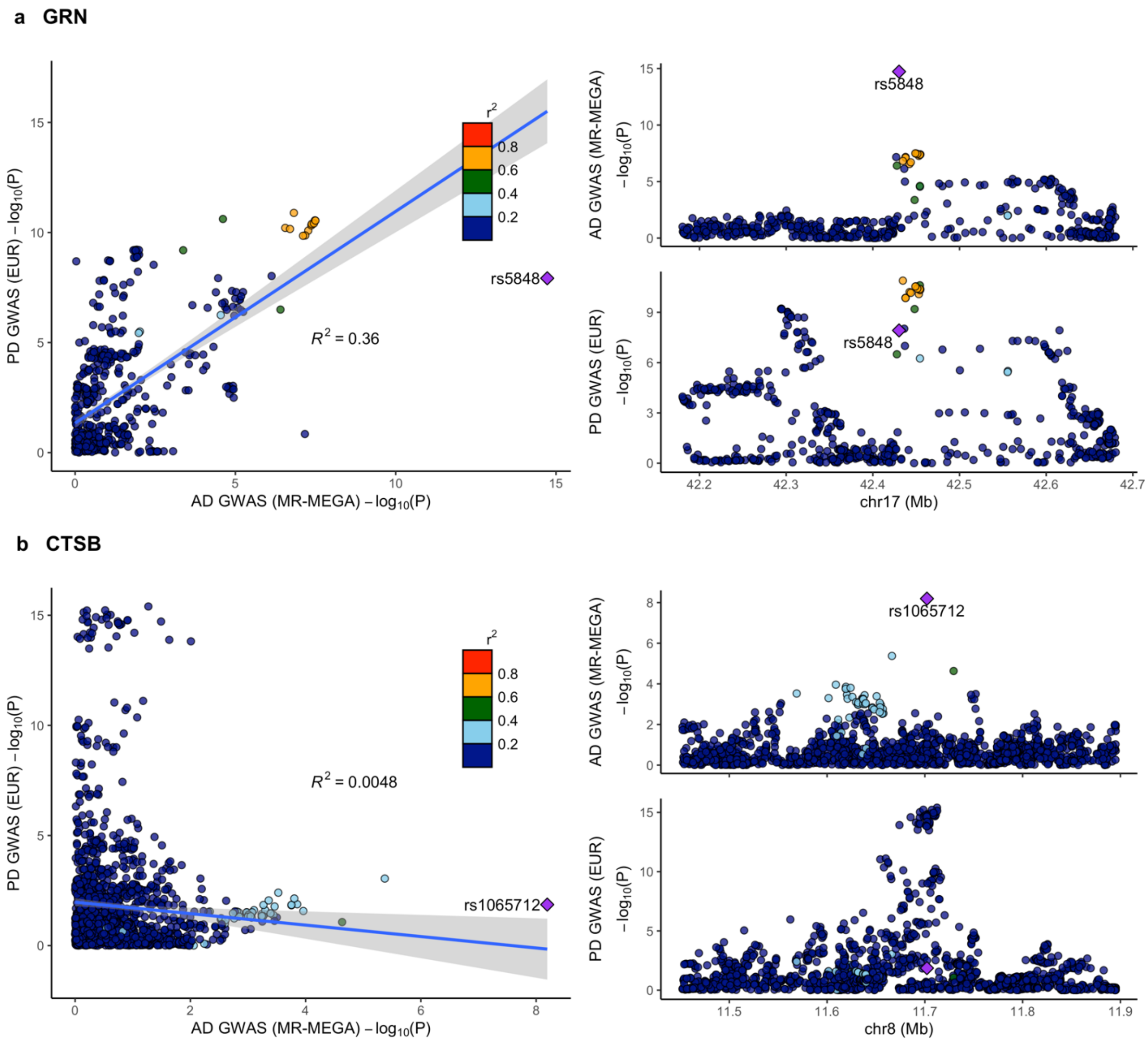


**Fig. S9:** LocusCompare plots for a) *GRN* and b) *CTSB* between AD and Parkinson's disease (PD). Reference LD patterns are based on the European population from 1000 Genomes. Points represent SNPs plotted at their -$\log_{10}$ P-valu

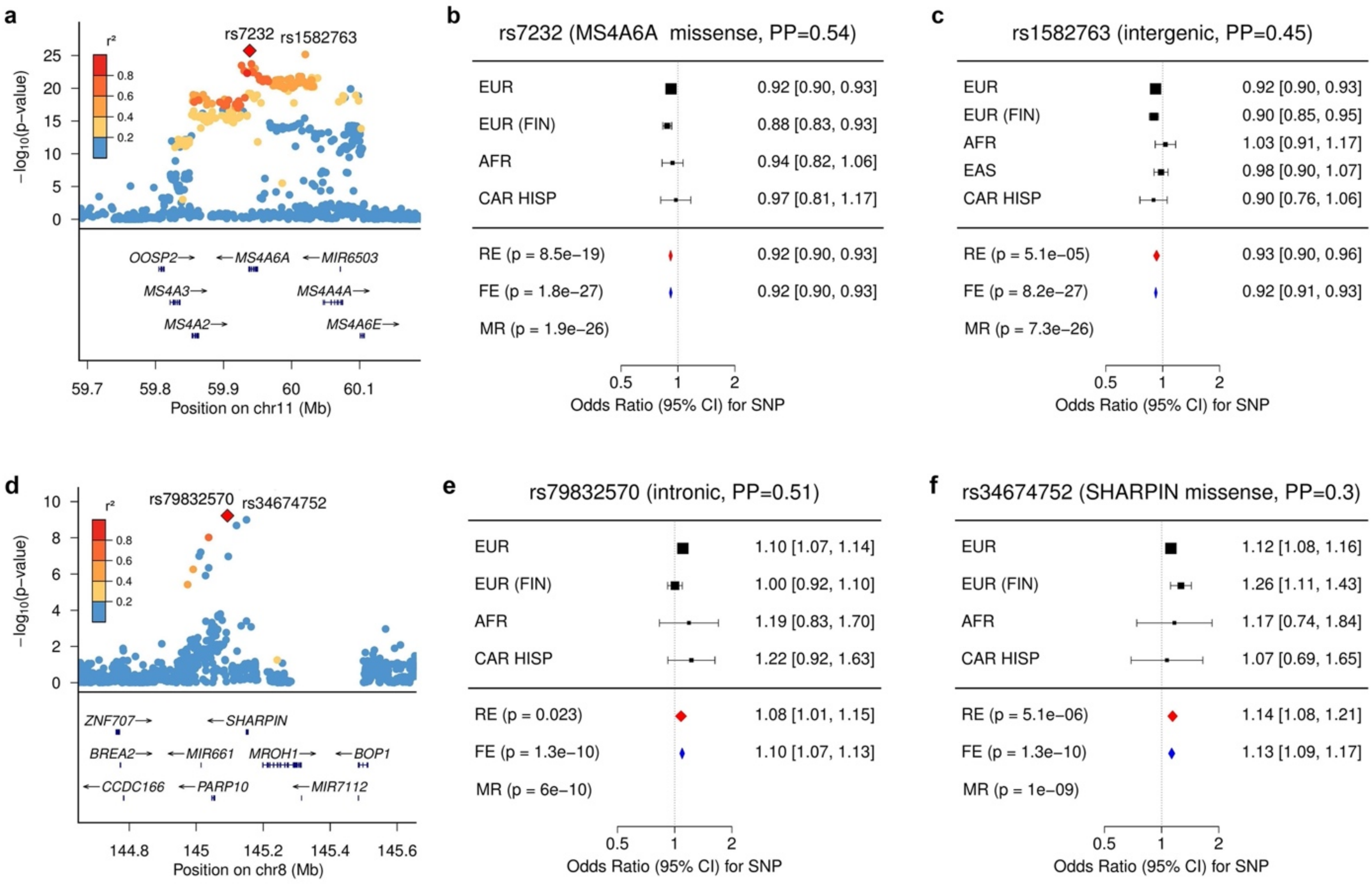


**Fig. S10:** Forest plot for fine-mapped missense variants in a) *MS4A6A*-rs7232 and d) *SHARPIN*-rs34674752 with complementary locus zooms in b) and f), respectively. Panels c) and e) show other top fine-mapped SNPs in these loci

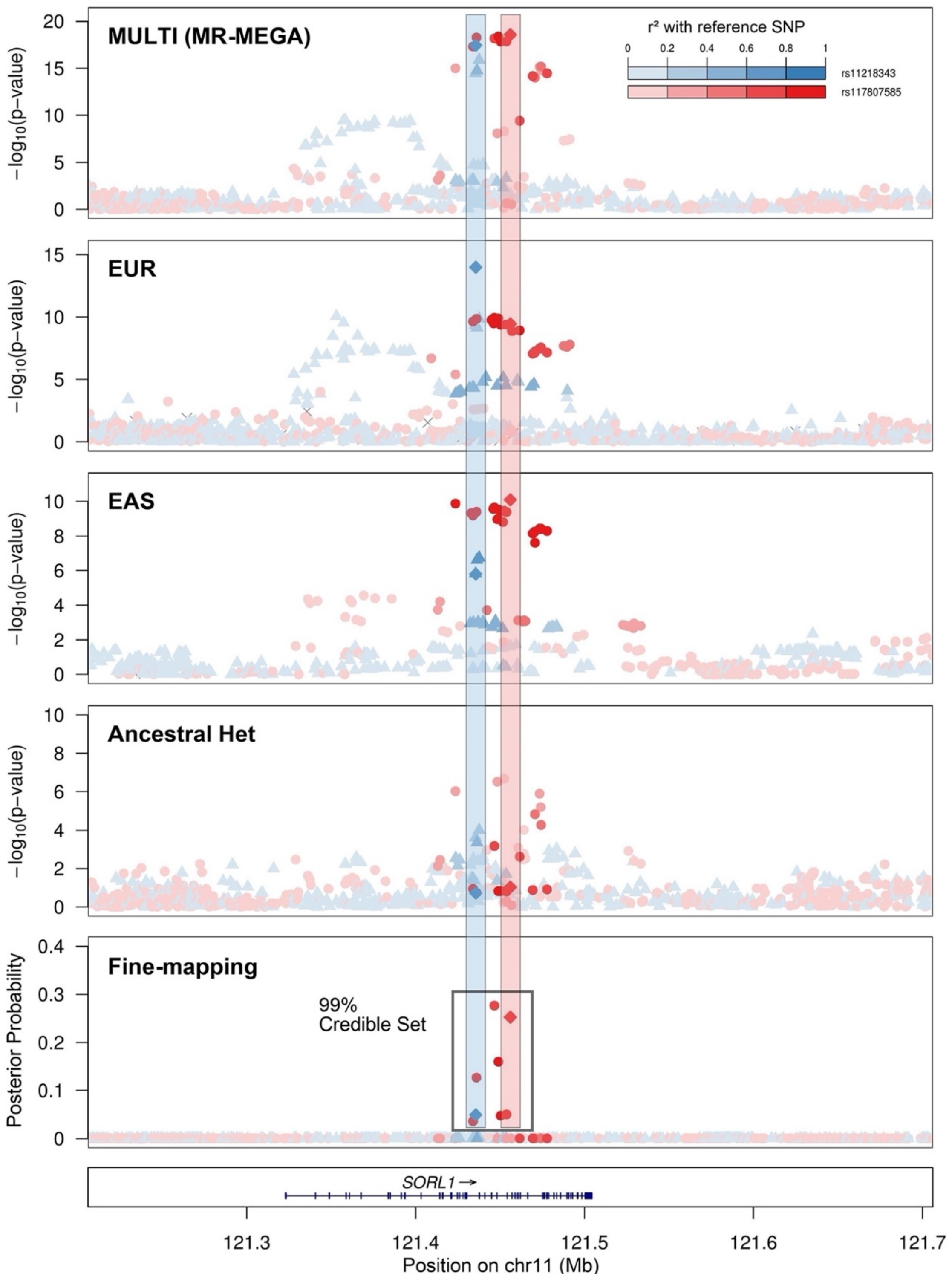


**Fig. S11:** LocusZoom plots showing different regional architecture at the *SORL1* locus in East Asian versus European populations. Diamond points represent the 2 different lead SNPs at this locus, *SORL1*-rs11218343 in Europeans and *SORL1*-rs117807585 in East Asians. SNPs are colored by LD with the respective population from 1000 Genomes.

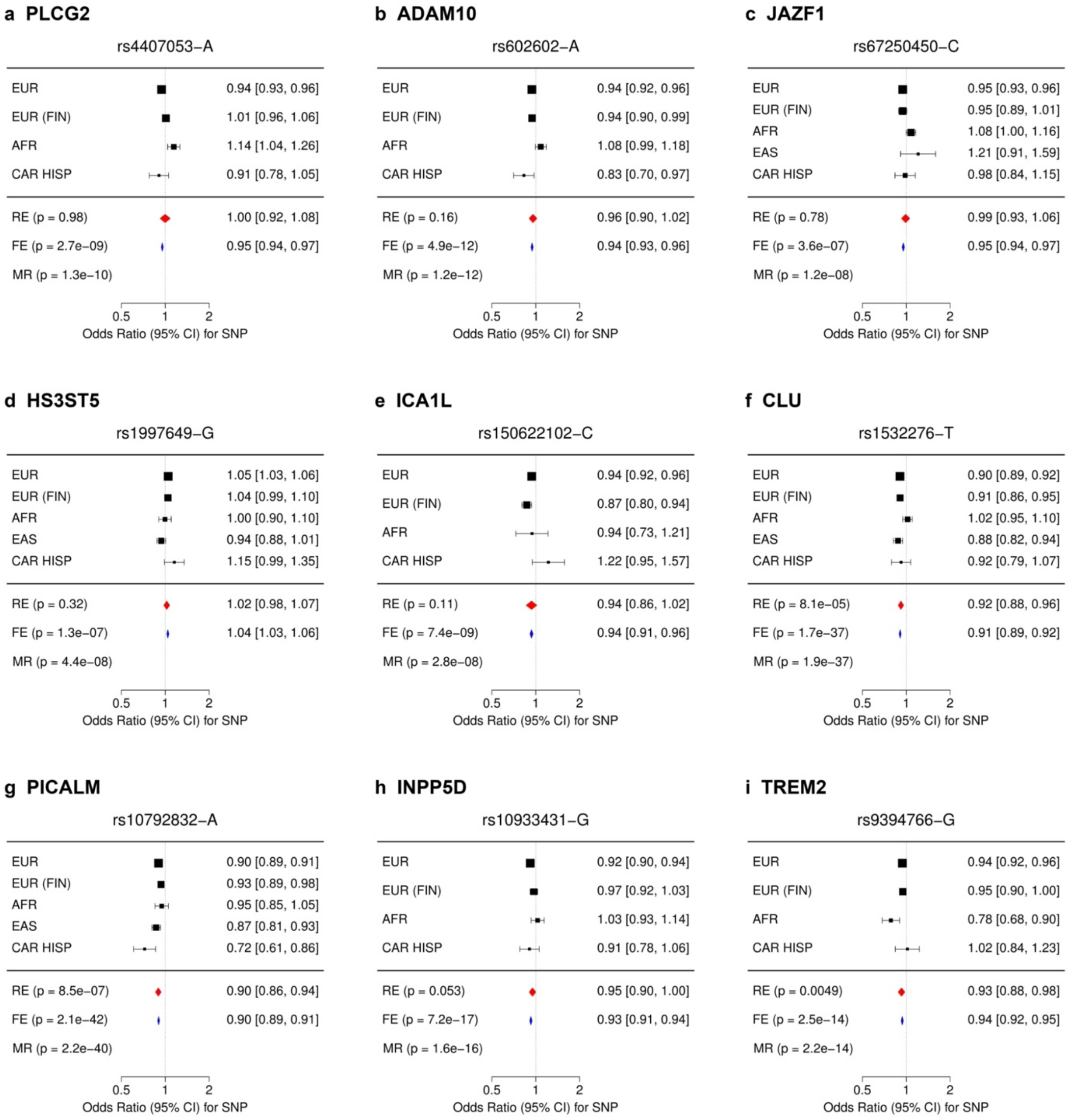
a PLCG2
rs4407053−A
EUR 0.94 [0.93, 0.96]
EUR (FIN) 1.01 [0.96, 1.06]
AFR 1.14 [1.04, 1.26]
CAR HISP 0.91 [0.78, 1.05]
RE (p = 0.98) 1.00 [0.92, 1.08]
FE (p = 2.7e−09) 0.95 [0.94, 0.97]
MR (p = 1.3e−10)
0.5 1 2
Odds Ratio (95% CI) for SNP
b ADAM10
rs602602−A
EUR 0.94 [0.92, 0.96]
EUR (FIN) 0.94 [0.90, 0.99]
AFR 1.08 [0.99, 1.18]
CAR HISP 0.83 [0.70, 0.97]
RE (p = 0.16) 0.96 [0.90, 1.02]
FE (p = 4.9e−12) 0.94 [0.93, 0.96]
MR (p = 1.2e−12)
0.5 1 2
Odds Ratio (95% CI) for SNP
c JAZF1
rs67250450−C
EUR 0.95 [0.93, 0.96]
EUR (FIN) 0.95 [0.89, 1.01]
AFR 1.08 [1.00, 1.16]
EAS 1.21 [0.91, 1.59]
CAR HISP 0.98 [0.84, 1.15]
RE (p = 0.78) 0.99 [0.93, 1.06]
FE (p = 3.6e−07) 0.95 [0.94, 0.97]
MR (p = 1.2e−08)
0.5 1 2
Odds Ratio (95% CI) for SNP
d HS3ST5
rs1997649−G
EUR 1.05 [1.03, 1.06]
EUR (FIN) 1.04 [0.99, 1.10]
AFR 1.00 [0.90, 1.10]
EAS 0.94 [0.88, 1.01]
CAR HISP 1.15 [0.99, 1.35]
RE (p = 0.32) 1.02 [0.98, 1.07]
FE (p = 1.3e−07) 1.04 [1.03, 1.06]
MR (p = 4.4e−08)
0.5 1 2
Odds Ratio (95% CI) for SNP
e ICA1L
rs150622102−C
EUR 0.94 [0.92, 0.96]
EUR (FIN) 0.87 [0.80, 0.94]
AFR 0.94 [0.73, 1.21]
CAR HISP 1.22 [0.95, 1.57]
RE (p = 0.11) 0.94 [0.86, 1.02]
FE (p = 7.4e−09) 0.94 [0.91, 0.96]
MR (p = 2.8e−08)
0.5 1 2
Odds Ratio (95% CI) for SNP
f CLU
rs1532276−T
EUR 0.90 [0.89, 0.92]
EUR (FIN) 0.91 [0.86, 0.95]
AFR 1.02 [0.95, 1.10]
EAS 0.88 [0.82, 0.94]
CAR HISP 0.92 [0.79, 1.07]
RE (p = 8.1e−05) 0.92 [0.88, 0.96]
FE (p = 1.7e−37) 0.91 [0.89, 0.92]
MR (p = 1.9e−37)
0.5 1 2
Odds Ratio (95% CI) for SNP
g PICALM
rs10792832−A
EUR 0.90 [0.89, 0.91]
EUR (FIN) 0.93 [0.89, 0.98]
AFR 0.95 [0.85, 1.05]
EAS 0.87 [0.81, 0.93]
CAR HISP 0.72 [0.61, 0.86]
RE (p = 8.5e−07) 0.90 [0.86, 0.94]
FE (p = 2.1e−42) 0.90 [0.89, 0.91]
MR (p = 2.2e−40)
0.5 1 2
Odds Ratio (95% CI) for SNP
h INPP5D
rs10933431−G
EUR 0.92 [0.90, 0.94]
EUR (FIN) 0.97 [0.92, 1.03]
AFR 1.03 [0.93, 1.14]
CAR HISP 0.91 [0.78, 1.06]
RE (p = 0.053) 0.95 [0.90, 1.00]
FE (p = 7.2e−17) 0.93 [0.91, 0.94]
MR (p = 1.6e−16)
0.5 1 2
Odds Ratio (95% CI) for SNP
i TREM2
rs9394766−G
EUR 0.94 [0.92, 0.96]
EUR (FIN) 0.95 [0.90, 1.00]
AFR 0.78 [0.68, 0.90]
CAR HISP 1.02 [0.84, 1.23]
RE (p = 0.0049) 0.93 [0.88, 0.98]
FE (p = 2.5e−14) 0.94 [0.92, 0.95]
MR (p = 2.2e−14)
0.5 1 2
Odds Ratio (95% CI) for SNP

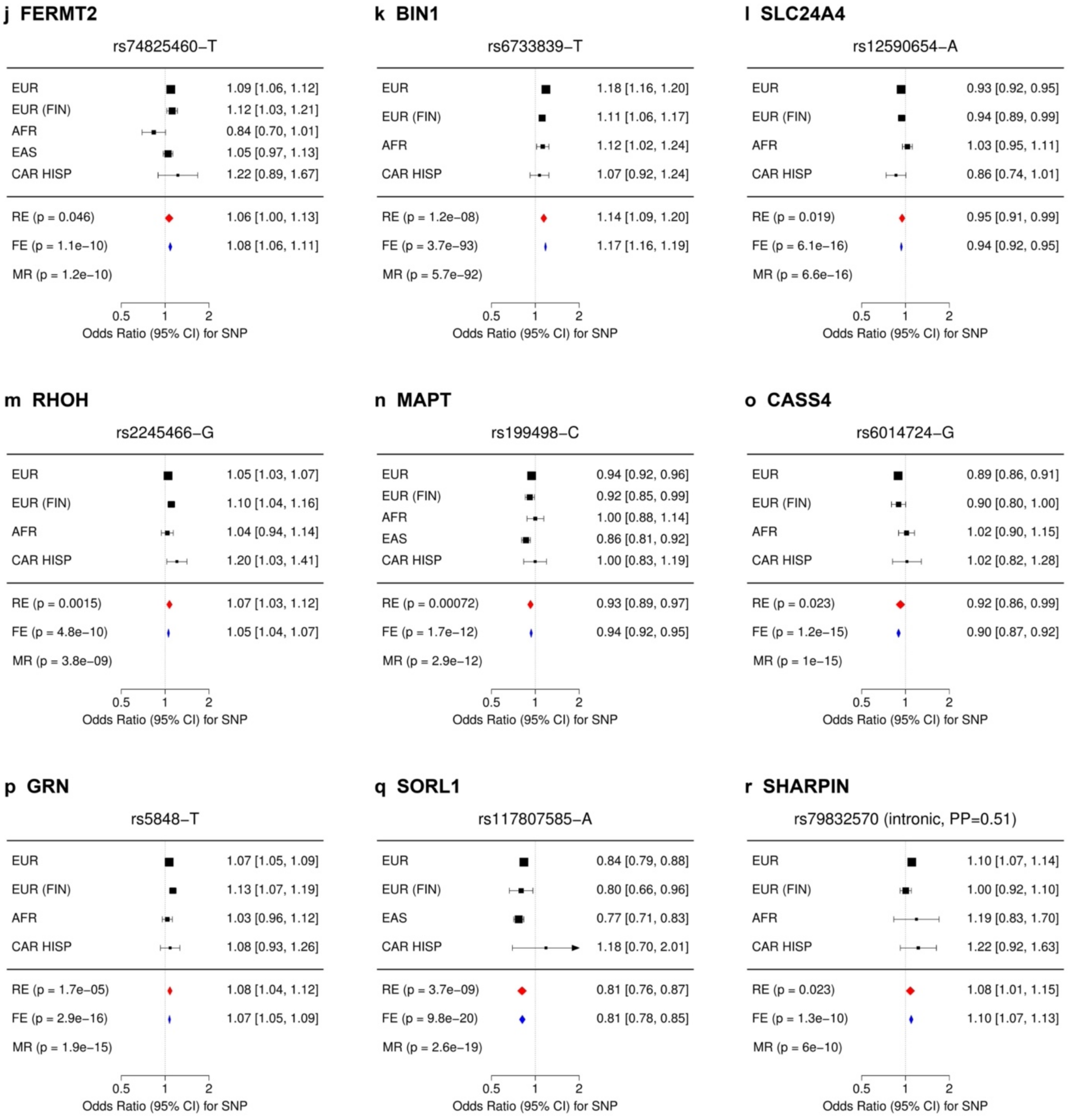


**Fig. S12:** Forest plots for loci with significant heterogeneity ($I^2$ > 30%) outside of the *APOE* region.

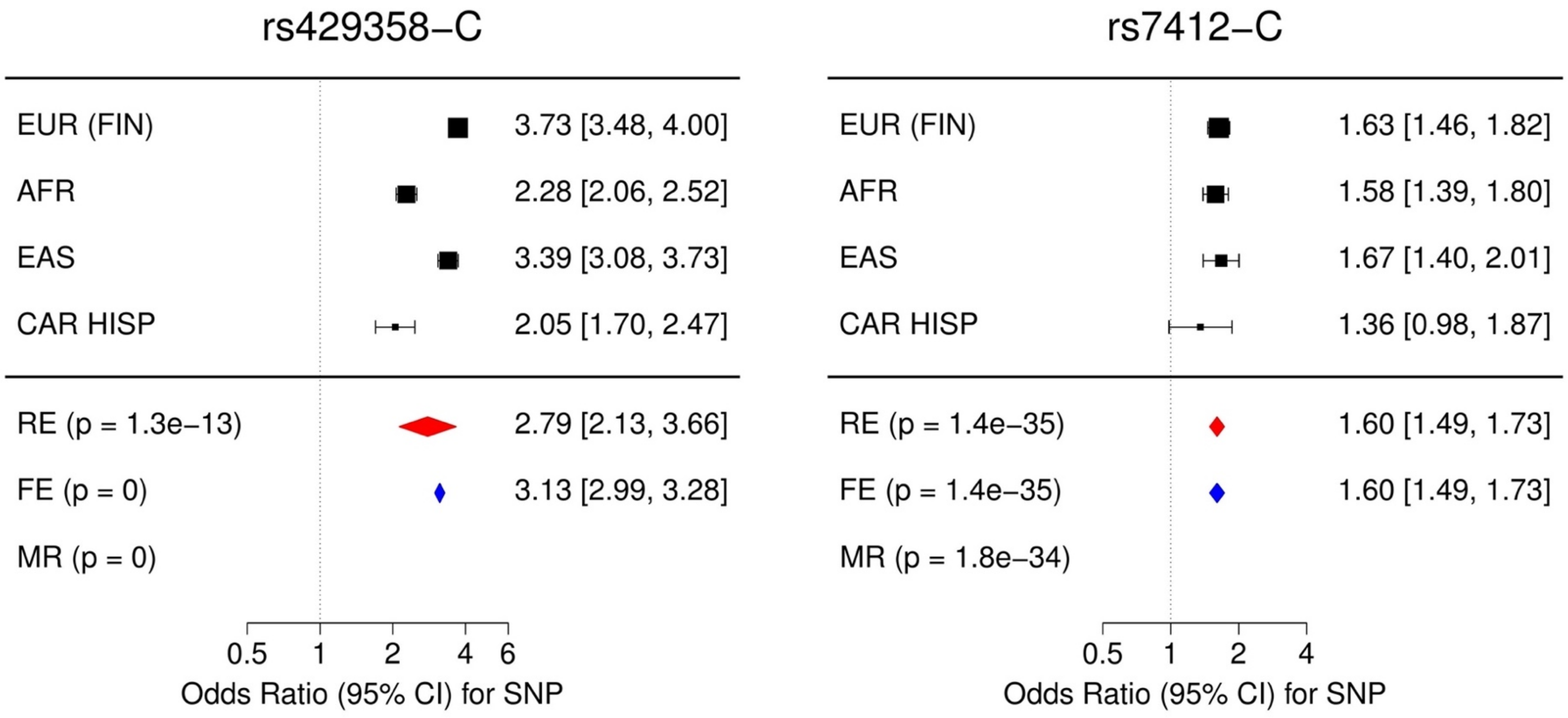


**Fig. S13:** Forest plots for *APOE*-rs429358 and *APOE*-rs7412.

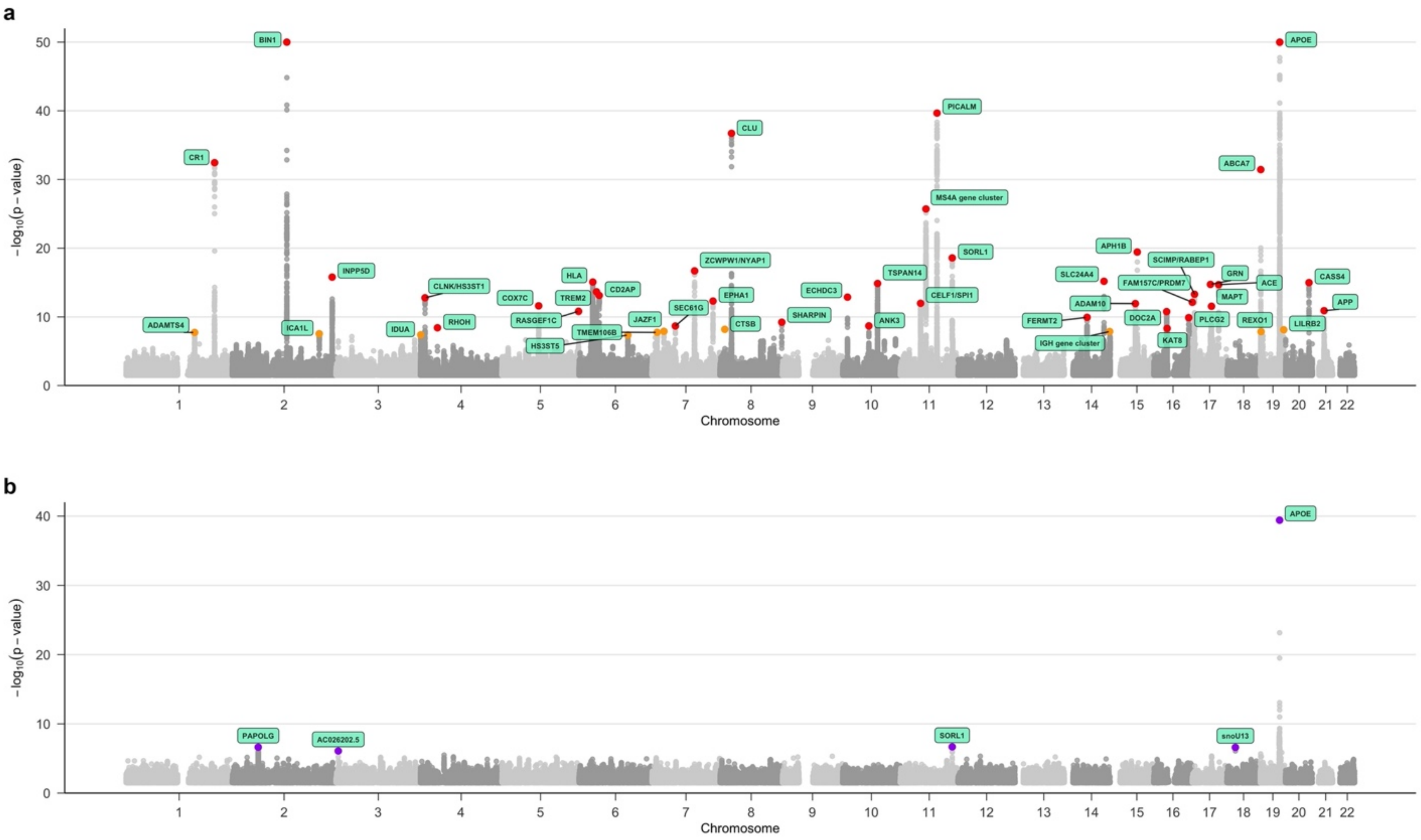


**Fig. S14:** a) Manhattan plot for MR-MEGA meta-analysis truncated at $-\log_{10}(P) < 50$. Red labeling corresponds to significance at $P < 5 \times 10^{-9}$ and orange corresponds to significance at $P < 5 \times 10^{-8}$. b) Manhattan plot for $P_{HET}$ from MR-MEGA truncated at $-\log_{10}(P) < 40$. Purple labeling corresponds to $P < 1 \times 10^{-6}$.

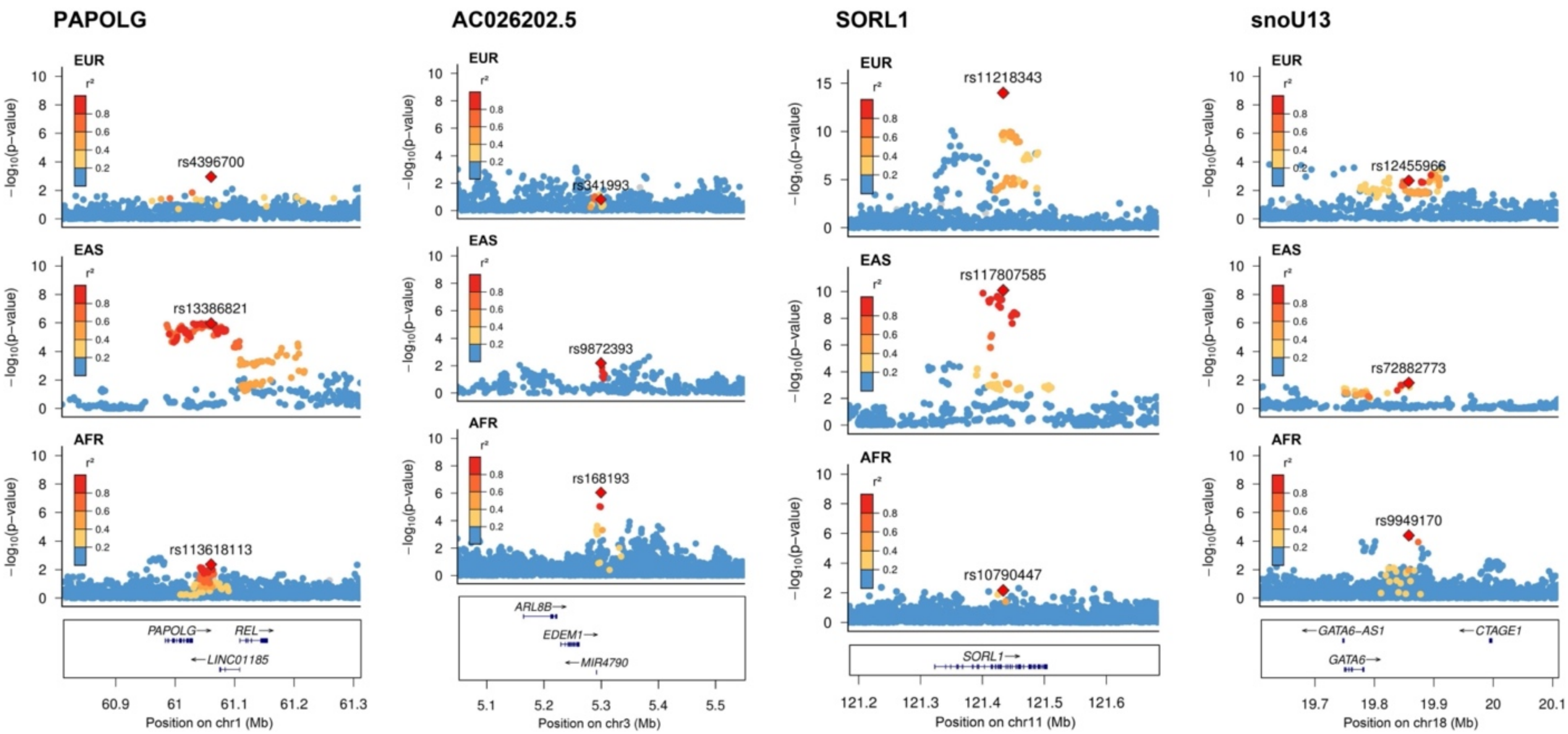


**Fig. S15a:** LocusZoom plots of loci showing significant ancestry-related heterogeneity ($P_{HET} < 1 \times 10^{-6}$) near *SORL1*, *PAPOLG*, *AC026202.5*, and *snoU13*. Labeled red diamonds correspond to the lead SNP in each ancestry group.

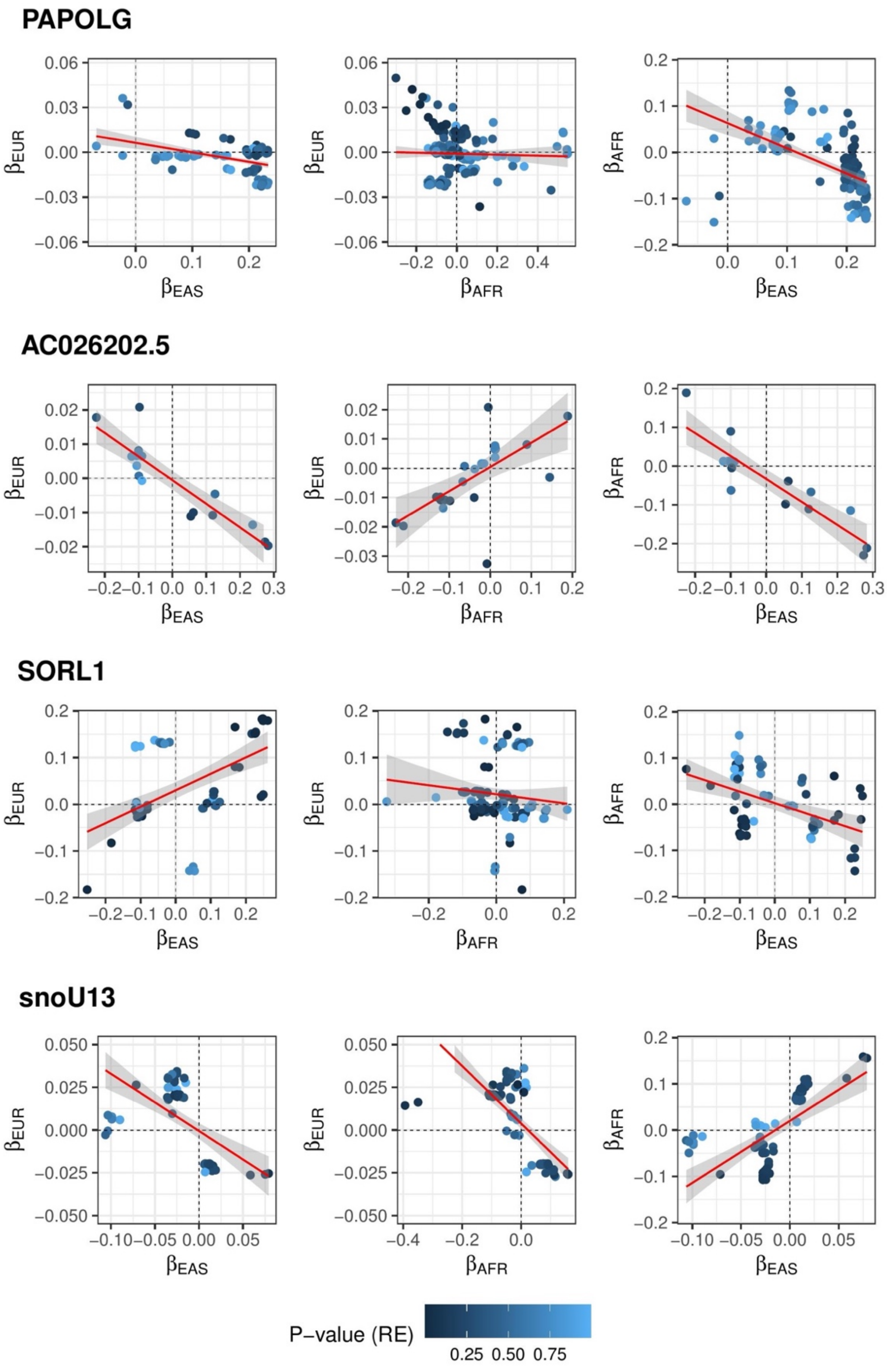


**Fig. S15b:** Beta-beta plots showing effect size correlation of *SORL1*, *PAPOLG*, *AC026202.5*, and *snoU13* across ancestry groups.

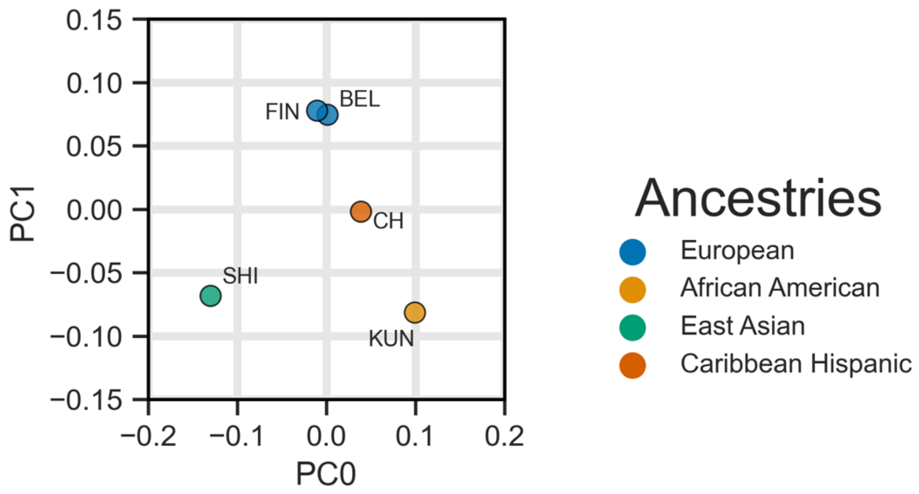


**Fig. S16:** The first 2 ancestral principal components (PCs) created and used by MR-MEGA plotted against each other, labeled by dataset (FIN: FinnGen R6, BEL: Bellenguez et al., CH: Caribbean Hispanic, SHI: Shigemizu et al., KUN: Kunkle et al.) and color coded by ancestry group.
